# Large language models for self-administered conversational vignette assessment of provider competencies: A pilot and validation study in Vietnam with automated LLM-powered transcript classification

**DOI:** 10.64898/2026.03.02.26347479

**Authors:** Benjamin Daniels, Wenjia(Stella) Zhang, Hoang Nguyen, David Duong

## Abstract

We developed and validated a self-administered clinical vignette method powered by a large language model (LLM), deployed through a web survey, to measure primary health care provider competencies in Vietnam. In a focus group, 9 physicians first tested the cases and rated their overall realism at 3.78/5 (95% CI 3.65 to 3.90). In the validation phase, 22 providers completed 132 vignette interactions across ten clinical scenarios in Vietnamese, of which 130 were coded by the research team and rated by expert clinicians. Essential diagnostic checklist scores correlated with the ratings of the two expert clinicians who graded the transcripts, one working in Vietnamese and one in English (Pearson’s *ρ* = 0.55 and 0.60). We classified the transcripts against the same rubric with a speaker-attributed GPT-5.4 pipeline, which agreed with the human coding on 90.1% (95% CI 88.0 to 92.2) of items from translated English transcripts and 89.2% (95% CI 86.8 to 91.5) from the original Vietnamese, at AUROCs of 0.912 (95% CI 0.899 to 0.923) and 0.922 (95% CI 0.913 to 0.930). Both language differences are signed Vietnamese minus English. Vietnamese coding was better at the item level, by an AUROC difference of 0.0100 (95% CI 0.0032 to 0.0168), and the session-level correlation difference of −0.089 (95% CI −0.181 to 0.002) includes zero once we cluster on providers, so a separate translation step does not appear necessary for this application. LLM-driven conversational vignettes offer a low-cost and scalable method for assessing provider competencies in respondents’ local language while preserving the open-ended format critical to vignette validity and enabling flexible feature extraction from transcripts using grading rubrics. The method is open-source and designed for replication in other health system contexts.

## 1 Introduction

Human resources for health represent the majority of recurrent health expenditures in most countries, yet investments in primary health care education and training have not consistently translated into improved clinical competencies [1, 2, 3, 4, 5]. Clinical vignettes – open-ended roleplay interactions in which an enumerator presents a standardized case – have emerged as a primary method for measuring provider competencies across health systems [6, 7]. By eliciting each provider’s unprompted approach to diagnosis and management of simulated cases, vignettes measure the best possible standard of care that providers are capable of delivering [8]. However, traditional vignette surveys require two trained enumerators per interaction, significant travel budgets, and in-person coordination, making them expensive to deploy at scale or repeat at frequent intervals [6, 7]. Digital multiple-choice formats undermine validity because unprompted elicitation of knowledge is the foundation of the method [6].

Recent advances in large language models (LLMs) have created new possibilities for scalable clinical assessment. LLM-based virtual patients can simulate realistic patient-clinician dialogues and provide automated performance feedback [9], and a rapidly growing literature has explored their use in medical education [10]. However, most existing applications target medical student training in high-income settings, and few have been validated against established competency measures or deployed for health workforce assessment in low- and middle-income countries, where the need is greatest [11]. Automated scoring of clinical interactions using LLMs has shown promise but remains noisy, with studies reporting moderate agreement with human evaluators at best [12, 13]. A tool that combines LLM-based patient simulation with automated data extraction – validated against expert clinical judgment – could enable routine, low-cost competency measurement without in-person enumerators.

This study presents a new method for self-administered and automatically coded clinical vignettes using LLMs. We developed an LLM-based chatbot interface [14, 15] using web-based survey software compatible with mobile phones (**Fig 1**). In Vietnam, we piloted and validated this method with primary health care providers across ten clinical scenarios. The chatbot – powered by OpenAI (gpt-4.1-nano) – role-played clinical cases in Vietnamese via respondents’ personal mobile devices for 132 interactions. We also developed an automated data extraction pipeline using OpenAI’s GPT-5.4 model (gpt-5.4-2026-03-05) to classify provider performance from the resulting transcripts. We report results from (1) a focus group of 9 providers assessing usability and realism, and (2) a separate validation sample of 22 providers who completed 132 interactions, of which 130 were fully coded and rated, comparing human-coded, LLM-coded, and expert-graded competency measures.

**Fig 1:**
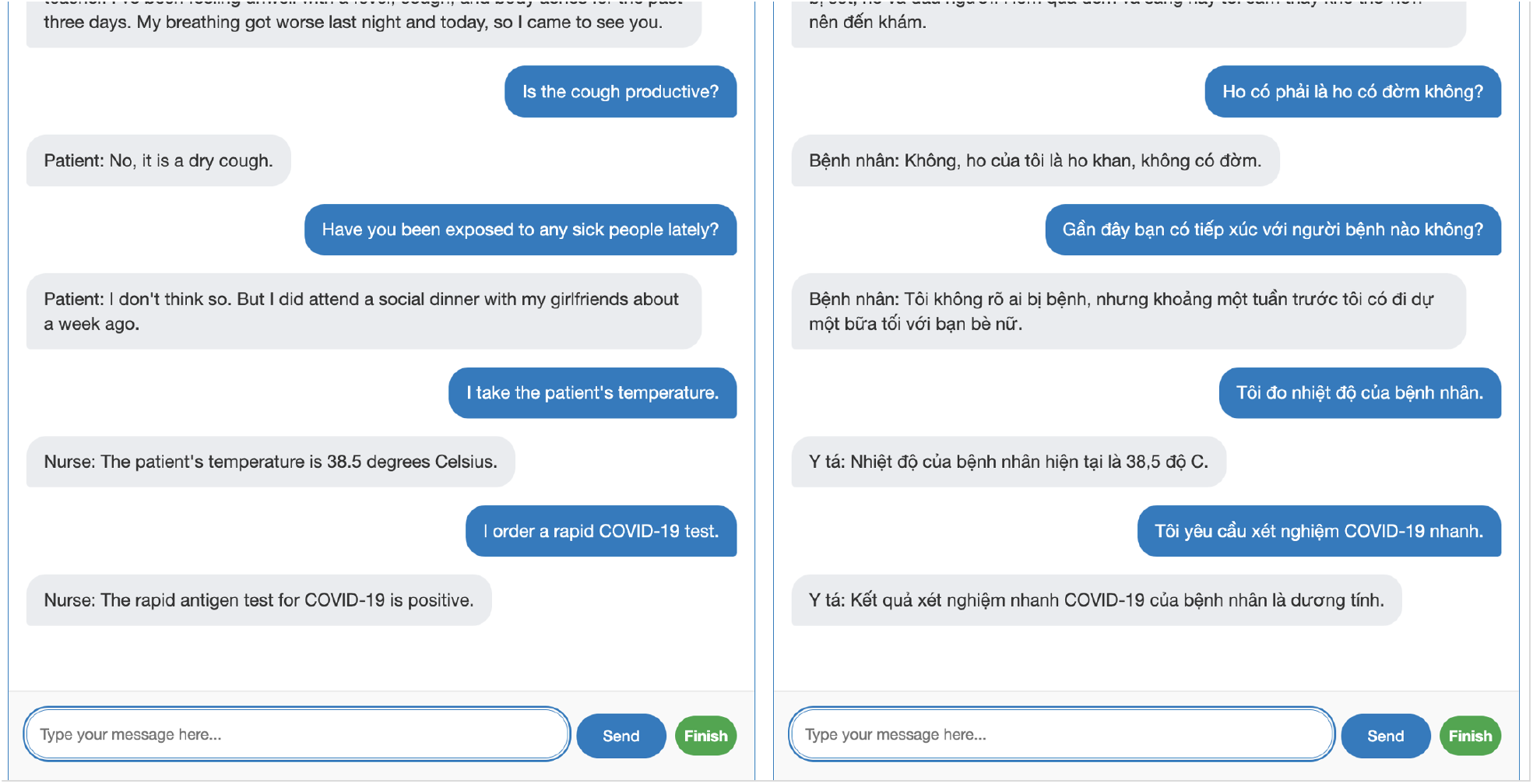
Screenshot of the LLM-based chatbot interface deployed through the web survey. Left: English translation. Right: Original Vietnamese. The provider (blue bubbles) asks clinical questions and orders examinations; the LLM-simulated patient (grey bubbles) responds according to a pre-scripted clinical scenario. The interface runs on any smartphone or laptop browser.

## 2 Materials and methods

### 2.1 Ethics Statement

All procedures were approved by the ethical review board of the Hanoi University of Public Health (025-375/DD-YTCC) and Mass General Brigham Institutional Review Board (Protocol Number 2023P003011). All participants provided written informed consent prior to participation.

### 2.2 Study design and participants

In August 2025, we administered a web-based survey (either by laptop or by mobile phone) that included LLM-based virtual patient “chatbots” preloaded with a pool of ten clinical cases (five general primary care conditions and five hepatitis-related) (**Table 1**). Each case scenario – detailing the patient’s socioeconomic background, chief complaint, medical history, clinical exam results, and potential responses to a set of “checklist” questions – was scripted by expert clinicians supporting the research team. The OpenAI application programming interface (API; model gpt-4.1-nano) was prompted through the survey software to perform according to these scenarios such that reasonable standardization was achieved across interactions. The scenarios developed for this study are available in the **Supporting Information**.

**Table 1:** Clinical case scenarios and validation summary. Each provider completed six randomly assigned cases (130 fully coded and rated interactions from 22 providers; 132 interactions fielded in total). Items column shows total checklist items per case (case-specific plus 19 common items administered across all cases); the overall row instead shows the 7814 rubric-item observations these interactions contain. Checklist completion and expert ratings are condition-level means.

| Case | Condition | Category | N | Items | Checklist (%) | Expert (VN) | Expert (EN) |
| --- | --- | --- | --- | --- | --- | --- | --- |
| 1 | Asthma | General | 14 | 62 | 12.1 | 3.07 | 3.71 |
| 2 | Pneumonia | General | 14 | 48 | 23.6 | 3.79 | 3.36 |
| 3 | Type II Diabetes Mellitus | General | 13 | 58 | 15.4 | 3.77 | 3.08 |
| 4 | Pulmonary Tuberculosis | General | 13 | 58 | 21.5 | 3.15 | 3.23 |
| 5 | Hypertension | General | 11 | 56 | 16.7 | 3.45 | 3.09 |
| 6 | Hepatitis B with Cirrhosis | Hepatitis | 14 | 74 | 11.4 | 3.07 | 2.50 |
| 7 | Hepatitis B in Pregnancy | Hepatitis | 12 | 61 | 13.3 | 3.42 | 3.17 |
| 8 | HBV Not Eligible for Treatment | Hepatitis | 13 | 58 | 15.5 | 3.46 | 3.15 |
| 9 | Hepatitis C | Hepatitis | 14 | 62 | 17.5 | 3.50 | 3.29 |
| 10 | Hepatitis C with ARV | Hepatitis | 12 | 63 | 16.6 | 3.50 | 3.50 |
| *Overall* |  |  | 130 | 7814 | 16.1 | 3.42 | 3.21 |

The research team first recruited and consented 9 physicians from a Provincial Hospital in Phu Tho, Vietnam, to a focus group for usability testing and UI/UX refinement. These providers received a live demonstration by the research team and were instructed to complete as many case scenarios as they found reasonable in a 45-minute period, followed by closed-ended (Likert scale) feedback questions on the realism of each case presentation. Two days later, the team conducted a structured group discussion to collect open-ended feedback, which was incorporated into the validation round.

For the validation sample, the research team contacted and consented 22 providers enrolled in the 2024-5 cohort of the StITCH hepatitis care training program, which co-designed and delivered a people-centered model of care for hepatitis B and C at the primary health care level in Vietnam [16]. The validation group was given a webinar orientation with no additional in-person support. These providers were assigned to complete six vignettes selected at random – three general cases and three hepatitis cases each. The validation fielding yielded 132 completed interactions. 2 of them were neither coded by the research team nor rated by the expert clinicians, so both the session-level and the item-level analyses reported below use the remaining 130 interactions. The fidelity audit of the chatbot covers all 132 fielded transcripts.

### 2.3 Chatbot implementation

The chatbot interface was built using web-based survey software with an LLM plugin [14, 15]. Each clinical case was encoded as a structured system prompt (1,100–1,500 words) containing: the patient’s demographic background and presenting complaint; a comprehensive medical and social history; pre-scripted responses to anticipated history-taking questions organized by clinical domain; clinical examination findings to be revealed when specific examinations were ordered; laboratory and imaging results for relevant tests; and the correct diagnosis with recommended treatment and counseling points. To maintain standardization, the system prompt instructed the model to report results as “normal” or “unremarkable” for tests not specified in the case scenario. The full set of case prompts is available in the **Supporting Information**.

The OpenAI model gpt-4.1-nano was selected after testing various model options for speed of response, cost-efficiency, and adequate performance in Vietnamese-language roleplay. The plugin called that model through the OpenAI API at the API’s default parameters. We report the fidelity audit as evidence for how the chatbot behaved. The survey plugin manages conversational state by appending each user message and model response to the conversation history, which is passed in full with each subsequent API call. Providers accessed the survey on their personal smartphones or laptops via a URL link. The chatbot responded in Vietnamese (selected by every provider) and maintained conversational context throughout each interaction. Providers could ask history-taking questions, order clinical examinations and laboratory tests, and provide a diagnosis and treatment plan, all through natural-language text input.

### 2.4 Data extraction and coding

Interaction transcripts were translated from Vietnamese to English using the Google Translate API and manually coded by the research team to determine whether each provider: (1) asked each of the pre-specified checklist history-taking questions, (2) ordered the recommended clinical examinations and laboratory tests, and (3) offered the correct diagnosis. A single coder produced this coding for every transcript, and no transcript was coded twice. We therefore report no inter-rater reliability statistic for the reference standard, and no procedure for resolving disagreements between coders was required. We also coded the same transcripts with GPT-5.4 (gpt-5.4-2026-03-05) at temperature zero, with JSON-mode output, response validation, and up to five retries on malformed responses. We ran both binary and confidence-scored coding on the original Vietnamese transcripts as well as the English translations, using the same rubric in both languages. Each language arm was scored once, in a single pass at temperature zero. Using the human-coded and LLM-coded checklist items separately, a checklist score was calculated for each vignette response as the proportion of items completed.

### 2.5 Chatbot fidelity audit

We audited every one of the 132 fielded transcripts against the case script, using the same GPT-5.4 configuration as the rubric coding and a separate prompt, reproduced in the **Supporting Information**. For each provider turn the audit recorded any examination or laboratory test the provider ordered and whether a result for that order was returned later in the conversation. We counted an order as returned when the chatbot supplied the scripted value for that test, and also when it gave the answer the system prompt directs for tests the script does not cover: *“If no specific test results are specified, give a normal range value or say the results are unremarkable or normal.”* It also recorded results the chatbot volunteered without being asked, turns delivered in a language other than the one the provider was writing in, breaks in the patient character, and statements contradicting the script. To detect drift in the scripted values of test results, we took each combination of case and test reported to at least three different providers and compared the numbers returned against the modal set for that combination.

### 2.6 Outcomes

For the focus group, primary outcomes were: (1) qualitative feedback on the LLM-based vignette survey obtained through the focus group discussion, and (2) quantitative ratings of scenario presentation realism. For the validation sample, the primary outcome was validation of the human-coded essential diagnostic checklist score via interaction-level correlations with expert clinician grading of the transcripts in English and in the original Vietnamese. Secondary outcomes were: (1) the reliability of the LLM-coded essential diagnostic checklist, assessed against human coding at the item and session levels; (2) a comparison of coding from translated English transcripts versus directly from the original Vietnamese transcripts; (3) case-level diagnostic accuracy and checklist completion; and (4) fidelity of the chatbot’s behavior to the case scripts.

### 2.7 Statistical analysis

For the focus group, the transcript was thematically analyzed by the research team to identify common themes for tool refinement. Provider perceptions of vignette realism were captured through structured survey questions rating overall realism, clinical presentation realism, and social/economic presentation realism. We calculated means and 95% confidence intervals for each.

For the validation sample, agreement between manual and automated data extraction was assessed by calculating the proportion of agreement, sensitivity, specificity, positive predictive value, and negative predictive value based on the human coding as ground truth, with true positives, false positives, true negatives, and false negatives reported by rubric domain. The discriminative ability of the LLM coding was evaluated using receiver operating characteristic (ROC) analysis: for each item, the LLM’s implied probability of positive classification – defined as the reported confidence when the binary score was 1 and one minus the confidence when the binary score was 0 – was used as a continuous predictor against the human-coded gold standard, and the area under the ROC curve (AUROC) was computed. Session-level correlations and regressions were estimated on the pooled sample with standard errors clustered at the provider level. Because each provider completed six vignettes, clustering was required even though no provider repeated the same case. We report the regressions in the direction shown in **Fig 2**, with human-coded checklist completion on the vertical axis. Differences between the English and Vietnamese codings were assessed with cluster-adjusted analytic tests: the item-level difference in AUROC as a difference in Somers’ D with a provider-clustered variance, and the session-level difference in correlations as a difference between two standardized regression slopes estimated jointly with a provider-clustered variance. Provider-cluster bootstrap intervals for both differences were computed as a robustness check, and the item-level difference was additionally tested with an exact provider-level sign-flip permutation test enumerating all 2^22^ sign assignments of the paired provider-level AUROC differences. The two expert clinicians who developed the vignettes scored provider performance on each transcript from 1–5 separately for history-taking, diagnosis accuracy, and management completeness, and we averaged those domain scores within rater to form the session-level expert ratings used here. The two raters worked independently and in parallel, each on their own schedule, and neither saw the other’s ratings, the checklist scores, or the research team’s coding of any transcript. One rater scored the original Vietnamese transcripts and the other scored the English translations, an assignment made during fielding to match each rater’s fluency and not prespecified in the design. Rater and language are therefore confounded, and the two expert columns cannot be interpreted as a comparison between the two languages.

**Fig 2:**
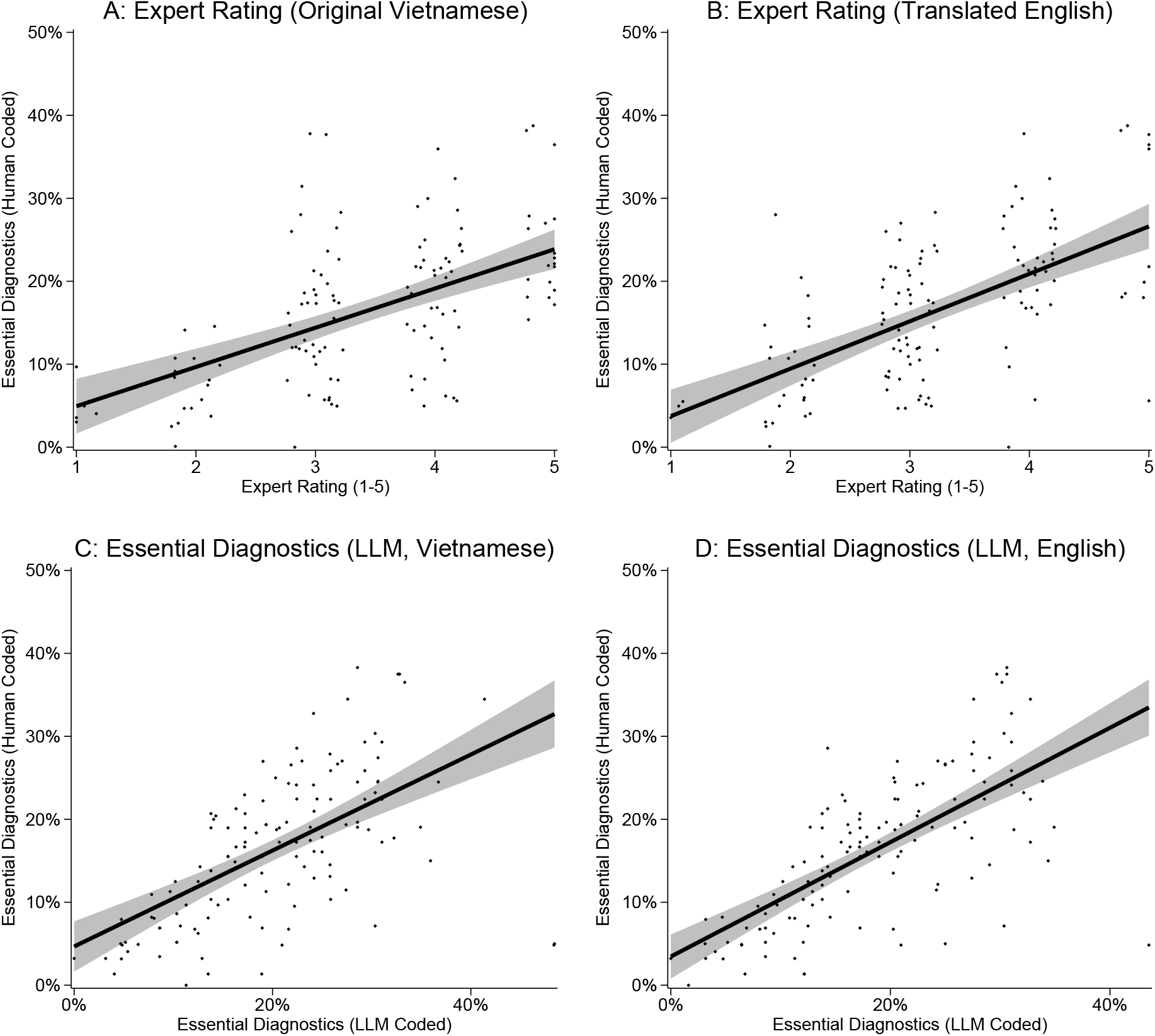
Validation of essential diagnostic checklist scores. Human-coded checklist completion (vertical axis) plotted against: Panel A: expert rating from original Vietnamese transcripts; Panel B: expert rating from translated English transcripts; Panel C: LLM-coded checklist directly from Vietnamese transcripts; Panel D: LLM-coded checklist from translated English transcripts. Each point is one provider–case interaction. Lines show OLS fit with 95% confidence interval.

This study is reported following the Strengthening the Reporting of Observational Studies in Epidemiology (STROBE) guidelines for the observational validation design. A completed checklist is provided in the **Supporting Information**.

### 3 Results

### 3.1 Focus group

Of the 9 providers, 7 completed six cases within the allotted 45-minute focus group session, with average interaction time ranging from 3.4 to 9.2 minutes per case. All interactions were conducted in Vietnamese on smartphones. Across all conditions, respondents rated overall case realism positively, at an average of 3.78/5 (95% CI 3.65 to 3.90).

Qualitative feedback highlighted several notable strengths. Providers consistently emphasized the ease of use and user-friendliness of the LLM-based conversational vignettes: *“Compared to other research tools, this one has a simpler interface and is easier to operate.”* Fast and contextually appropriate responses were the most frequently cited benefits (4 mentions), with providers noting that information provided was sufficient and meaningful for clinical decision-making: *“The virtual patient’s responses are relatively accurate, realistic, and easy to work with.”* Others commended its realism, including language localization and the ability to return to history-taking after laboratory results were delivered: *“The language has been fully localized into Vietnamese and [is] very understandable”*; *“It doesn’t give us everything at once for us to guess. The more we ask, [*… *] the more accurate the diagnosis will be.”* Another physician summarized: *“Regarding the system’s responses [*… *], the level of realism is good. If we guide or structure our questions clearly, it will deliver as expected.”*

A few respondents noted minor issues. The most common limitation was that certain laboratory or imaging tests were not performed or had results omitted (4 mentions). *“Some para-clinical tests weren’t done, but when I asked a few related questions, [the responses] felt complete.”* Although the limited test results raised concerns regarding realism, this was an intentional design feature to ensure reasonable standardization of the virtual patients: tests deemed irrelevant for the case were prompted to have results reported as “normal” or “unremarkable.” The absence of non-verbal cues was also considered a barrier to realistic clinical interaction: *“When we see patients in person, we can quickly evaluate whether there is an infectious syndrome. For asthma patients, just by observing their breathing we can tell how severe their respiratory distress is. But in a simulation, we are limited in the questions we can ask.”*

Respondents recommended several enhancements for future development. Suggestions included integrating visual elements such as laboratory images and video patient profiles, adding automatic performance feedback, and eventually transitioning from a web-based survey to an independent, offline smartphone application. One respondent also mentioned the potential benefit of including more complicated case scenarios to match clinical reality: *“In real hospitals, most patients don’t just have one disease – they usually have multiple comorbidities.”* Overall, the focus group participants judged the LLM-based vignette survey to be both usable and realistic.

### 3.2 Validation sample

22 providers completed 132 interactions in the validation sample, of which 130 were coded by the research team and rated by both expert clinicians. Those 130 interactions were the ones reported in **Table 1** and **Fig 2**. Providers completed 16.1% of checklist items on average across the 130 coded interactions (7814 item observations), reflecting the deliberately comprehensive nature of the rubrics. Each case included case-specific items plus 19 common items administered across all cases, so the benchmark was the full domain of possible actions rather than a minimum passing threshold. The expert ratings correlated with the human-coded checklist at 0.55 (95% CI 0.39 to 0.67) in Vietnamese and 0.60 (95% CI 0.38 to 0.74) in English. With human-coded checklist completion on the vertical axis, a one-point increase in the Vietnamese expert rating corresponded to 4.73 (95% CI 3.79 to 5.66, *R*^2^ = 0.30) higher checklist completion, and a one-point increase in the English expert rating corresponded to 5.72 (95% CI 3.91 to 7.53, *R*^2^ = 0.36) higher checklist completion.

The automated coding agreed with the human coding on most rubric items. From translated English transcripts, overall agreement was 90.1% (95% CI 88.0 to 92.2), with sensitivity 77.0% (95% CI 74.4 to 79.5), specificity 92.6% (95% CI 90.3 to 94.8), positive predictive value 66.5% (95% CI 58.1 to 74.9), and negative predictive value 95.5% (95% CI 94.6 to 96.3). The overall completion rate was 16.1% (95% CI 13.3% to 18.8%), so most item observations were true negatives and the positive predictive value depended chiefly on the specificity. At the session level, the English-coded checklist correlated with the human-coded checklist at 0.68 (95% CI 0.51 to 0.83), and the regression shown in **Fig 2** implied 0.69 (95% CI 0.46 to 0.92, *R*^2^ = 0.47) higher human-coded completion for a one-unit increase in the LLM-coded score. A one-unit increase in the human-coded score predicted 0.68 (95% CI 0.53 to 0.83) higher LLM-coded completion, and the slope remained below one, consistent with compression of high-performing sessions.

Direct coding from the original Vietnamese transcripts gave nearly the same operating characteristics. Overall agreement was 89.2% (95% CI 86.8 to 91.5), with sensitivity 78.8% (95% CI 77.1 to 80.5), specificity 91.1% (95% CI 88.4 to 93.9), positive predictive value 63.0% (95% CI 54.2 to 71.8), and negative predictive value 95.7% (95% CI 94.8 to 96.6). Sensitivity in the Vietnamese arm was therefore 78.8% against 77.0% in the English arm, at specificities of 91.1% and 92.6%. The session-level correlation with human coding was 0.60 (95% CI 0.38 to 0.80), against 0.68 in English, and the corresponding regression in **Fig 2** was 0.58 (95% CI 0.26 to 0.90, *R*^2^ = 0.35). A one-unit increase in the human-coded score predicted 0.61 (95% CI 0.41 to 0.81) higher LLM-coded completion, again below one. The two LLM codings remained highly correlated with each other, at Pearson’s *ρ* = 0.87 (95% CI 0.83 to 0.91).

Treating the confidence-weighted LLM output as a continuous classifier, the AUROC was 0.912 (95% CI 0.899 to 0.923) for English transcript coding and 0.922 (95% CI 0.913 to 0.930) for Vietnamese transcript coding (**Fig 3**). We report both language differences as Vietnamese minus English. The session-level difference in correlation was −0.089 (95% CI −0.181 to 0.002), *z* = −1.91, *p* = 0.056. At the item level the difference in AUROC was 0.0100 (95% CI 0.0032 to 0.0168), *z* = 2.87, *p* = 0.004, and the Vietnamese coding discriminated better; the exact permutation test agreed (*p* = 0.0013, on an unweighted provider-mean difference of 0.0131). Taken together, we detected no session-level difference between the two languages, the Vietnamese coding was modestly better at the item level, so a separate translation step is not required for automated checklist coding in this setting.

**Fig 3:**
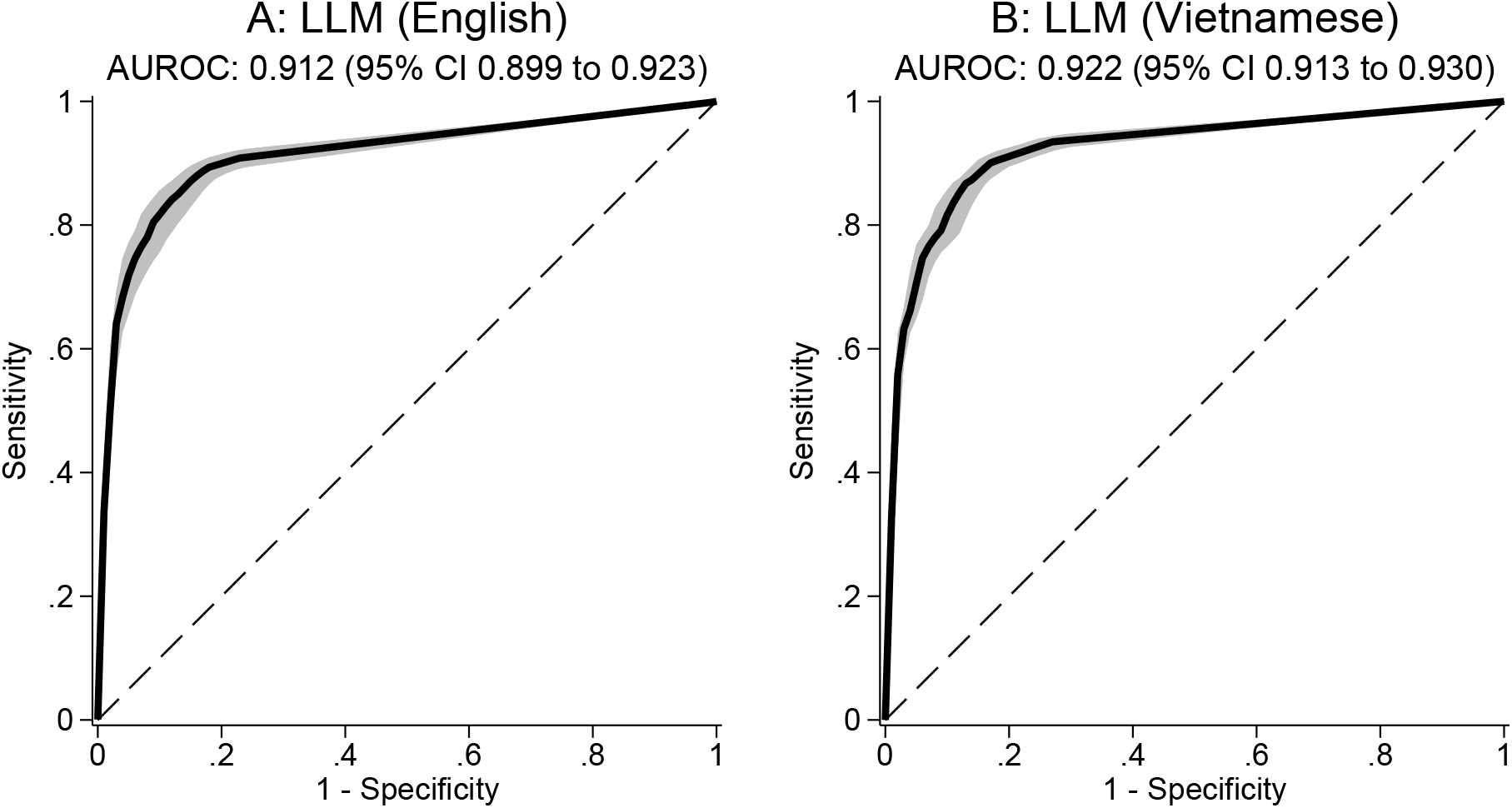
Receiver operating characteristic (ROC) curves for LLM coding of checklist items against human-coded gold standard. Panel A: coding from translated English transcripts. Panel B: coding directly from Vietnamese transcripts. Solid lines are the full-sample empirical ROC curves. Shaded areas are pointwise 95% confidence bands from a provider-cluster bootstrap, taking the 2.5th and 97.5th percentiles of the resampled sensitivity at each false-positive rate. The diagonal dashed line represents chance performance (AUROC = 0.5). Subtitles report the AUROC with its provider-clustered 95% confidence interval.

Automated scoring errors were not spread evenly across the rubric. **Table 2** reports agreement for each rubric domain. Laboratory testing produced 120 false positives across 816 observations, the highest rate of false credit anywhere in the rubric, at an agreement of 81.4%. Focus-group participants had flagged that same domain, noting that the chatbot sometimes omitted laboratory results, and the fidelity audit reported below found the chatbot’s own weakness in the same place. Testing items were the weakest domain, at 74.5% across 415 observations, on 76 false negatives against only 30 false positives. History taking was the largest domain at 2775 observations, with agreement of 90.9%, and the 19 common items administered in every case were the most reliably coded, at 95.2%.

**Table 2:** Agreement of LLM coding with human coding, by rubric domain, English arm. Agreement is the proportion of item observations on which the LLM coding and the human coding agreed, computed from translated English transcripts. Sensitivity is the proportion of human-coded positive items the LLM coding also scored positive, and specificity the proportion of human-coded negative items it also scored negative. True positives, false positives, true negatives, and false negatives are counts against the human coding as ground truth. Common items are the 19 rubric items administered in every case.

| Rubric domain | Observations | Agreement | Sensitivity | Specificity | True positives | False positives | True negatives | False negatives |
| --- | --- | --- | --- | --- | --- | --- | --- | --- |
| History taking | 2775 | 90.9% | 90.6% | 90.9% | 425 | 209 | 2097 | 44 |
| Clinical examination | 354 | 92.1% | 93.3% | 92.0% | 14 | 27 | 312 | 1 |
| Testing | 415 | 74.5% | 31.5% | 90.1% | 35 | 30 | 274 | 76 |
| Laboratory testing | 816 | 81.4% | 90.6% | 74.7% | 310 | 120 | 354 | 32 |
| Management | 250 | 89.2% | 13.8% | 99.1% | 4 | 2 | 219 | 25 |
| Counseling | 544 | 85.1% | 40.9% | 94.2% | 38 | 26 | 425 | 55 |
| Common items | 2660 | 95.2% | 71.4% | 97.0% | 140 | 73 | 2391 | 56 |

**Table 3** reports checklist completion and diagnostic accuracy for each case, all as pooled proportions. Checklist completion counted rubric items, which we report with provider-cluster bootstrap percentile intervals. Per-case diagnosis and qualifier proportions report Clopper-Pearson exact intervals. Correct diagnosis reports whether the provider named the condition. Across the five general cases, providers gave the correct diagnosis in 83.1% (95% CI 73.6 to 90.9) of 65 interactions; across the five hepatitis cases, in 92.3% (95% CI

**Table 3:** Checklist completion and diagnostic accuracy by case, with provider-cluster bootstrap and exact binomial intervals. *N* is the number of interactions for that case and Items is the number of checklist items in that case; in the overall row, Items is instead the 7814 rubric-item observations pooled across cases. Checklist completion is the pooled item-level proportion. Correct diagnosis is the proportion of interactions in which the provider named the condition, which scores all ten cases the same way. Qualifier identified is the proportion in which the provider also gave the stage or qualification that case’s rule specifies; only the five hepatitis rules specify one, so the five general cases report no value. Intervals are provider-cluster bootstrap percentiles for checklist completion and for the pooled diagnosis and qualifier proportions in the overall row, and Clopper-Pearson exact for the per-case diagnosis and qualifier proportions, all at the 95% level.

| Condition | $N$ | Items | Checklist (%) | Correct diagnosis (%) | Qualifier identified (%) |
| --- | --- | --- | --- | --- | --- |
| Asthma | 14 | 62 | 12.1 ( 8.6, 15.8) | 64.3 (35.1, 87.2) | – |
| Pneumonia | 14 | 48 | 23.6 (20.1, 27.6) | 92.9 (66.1, 99.8) | – |
| Type II Diabetes Mellitus | 13 | 58 | 15.4 (10.7, 20.4) | 84.6 (54.6, 98.1) | – |
| Pulmonary Tuberculosis | 13 | 58 | 21.5 (17.3, 25.7) | 92.3 (64.0, 99.8) | – |
| Hypertension | 11 | 56 | 16.7 (13.1, 20.8) | 81.8 (48.2, 97.7) | – |
| Hepatitis B with Cirrhosis | 14 | 74 | 11.4 ( 7.1, 16.1) | 92.9 (66.1, 99.8) | 42.9 (17.7, 71.1) |
| Hepatitis B in Pregnancy | 12 | 61 | 13.3 ( 8.6, 18.2) | 91.7 (61.5, 99.8) | 16.7 ( 2.1, 48.4) |
| HBV Not Eligible for Treatment | 13 | 58 | 15.5 (12.1, 19.4) | 100.0 (75.3, 100.0) | 0.0 ( 0.0, 24.7) |
| Hepatitis C | 14 | 62 | 17.5 (12.6, 22.5) | 92.9 (66.1, 99.8) | 14.3 ( 1.8, 42.8) |
| Hepatitis C with ARV | 12 | 63 | 16.6 (11.5, 22.0) | 83.3 (51.6, 97.9) | 33.3 ( 9.9, 65.1) |
| *Overall* | 130 | 7814 | 16.1 (13.3, 18.8) | 87.7 (79.7, 93.9) | 21.5 (12.1, 30.8) |

81.5 to 100.0) of 65. Providers named the condition in 87.7% (95% CI 79.7 to 93.9) of 130 interactions. The general items accepted a one-word answer, asking only whether the provider correctly diagnosed asthma, or pneumonia, or hypertension. Each hepatitis item also asked for a specific stage or qualification: hepatitis B with decompensated cirrhosis at Child-Pugh Class C in one case, chronic hepatitis C with advanced fibrosis at F3 without cirrhosis and without co-infection in another, chronic viremic hepatitis C co-infected with hepatitis B and HIV with compensated cirrhosis in a third, and chronic hepatitis B without fibrosis in the remaining two. The second diagnosis column reports that qualification, and providers gave it in 21.5% (95% CI 12.1 to 30.8) of 65 interactions.

### 3.3 Chatbot fidelity

We audited all 132 transcripts against the case scripts. Providers issued 840 orders for examinations or laboratory tests across those interactions. Of those, 112 never produced a result, 13.3% (95% CI 11.1% to 15.8%) of all orders, and 34 interactions contained at least one, 25.8% (95% CI 18.5% to 34.1%). The ten worst-affected interactions accounted for 70 of the 112 unanswered orders, or 62% of them. In the Hepatitis B in Pregnancy case, a provider asked *“Because hepatitis B can be transmitted from mother to child, you will have some tests done to decide whether you need to take medication to prevent transmission to your child, including the following tests: Blood count, blood biochemistry: creatinine, AST, ALT, HBV DNA, Anti HCV, Anti HIV, fetal ultrasound.”* and the chatbot answered *“[Nurse] Yes, I will take blood to do blood count, blood biochemistry (creatinine, AST, ALT), HBV DNA, Anti HCV, Anti HIV and perform fetal ultrasound for Ms. Pham. Please wait a moment for the sample to be taken and tested.”*; no value followed in the rest of the conversation. The case script supplied values for several of the tests ordered: *“AST/ALT: 19/20 U/L; HBeAg: Negative, HBV DNA: 200 IU/mL; Anti-HCV and HIV: Negative”*.

No turn was delivered in the wrong language. Where the same scripted value was reported to at least three different providers, 2 of 281 reported values were discordant with the modal set for that case and test, 0.7% (95% CI 0.1% to 2.5%). In the opposite direction from the missing results, the chatbot volunteered findings that nothing the provider asked for could plausibly cover in 39 interactions, 29.5% (95% CI 21.9% to 38.1%). In the Type II Diabetes Mellitus case, a provider asked *“How much is the patient’s blood sugar?”* and the chatbot returned *“[Nurse] The results of the patient’s blood sugar test are as follows: - Fasting blood sugar: 12.8 mmol/L - Random blood sugar: 16.7 mmol/L - HbA1c: 6.9% [Patient] My blood sugar seems high, doctor.”*.

## 4 Discussion

We developed and validated an LLM-based vignette methodology that can field conversational clinical cases remotely, preserve the open-ended structure that gives vignettes their validity, and produce checklist-based measures that align with expert ratings. The expert-session correlations were 0.55 and 0.60, and 132 clinical interactions were completed. The practical attraction of the method is that it replaces in-person enumeration with a respondent’s own phone or laptop while retaining the structure of a clinical conversation [6]. LLM-graded scores following the same human audits produced a correlated score consistent with a noisy interpretation of checklist item completeness. Regressing the LLM-coded score on the human-coded score gives a slope of 0.68 (95% CI 0.53 to 0.83) in English and 0.61 (95% CI 0.41 to 0.81) in Vietnamese. Measurement error in the human coding would also produce a slope below one, so these data do not tell us whether the classifier compresses the scale or the reference standard is noisy.

Our approach extends the emerging literature on LLM-based virtual patients [10, 9] in three ways. First, whereas most existing applications target medical students in high-income educational settings, we demonstrate feasibility for health workforce assessment among practicing providers in a low- and middle-income country context. Second, we validate the resulting competency scores against expert clinician ratings produced from the transcripts themselves, while recognizing that these raters were part of vignette development. Third, direct Vietnamese coding is sufficient for this use case. Item-level discrimination is better in Vietnamese by 0.0100 (95% CI 0.0032 to 0.0168) in AUROC, an interval that excludes zero, and the session-level correlation difference, signed the same way, is −0.089 (95% CI −0.181 to 0.002) and includes zero once we cluster on providers. Translation therefore appears unnecessary, though not literally equivalent, for automated checklist coding in this setting. Future directions for tool refinement may include incorporating audio or video patient profiles to enhance realism through non-verbal cues.

The need for scalable competency measurement is acute. Evidence from sub-Saharan Africa [3], South and Southeast Asia [17, 18, 19, 20], and Vietnam specifically [21, 22] demonstrates that delivered care quality remains low despite growing investments in medical education, and that progress has been slow. The most recent comprehensive assessment of primary care quality in Vietnam using clinical vignettes was conducted in 2015 [21]; the expense of replication has prevented a post-pandemic update. Methods like the one presented here could enable continuous, routine monitoring at a fraction of the cost of traditional approaches.

### 4.1 Limitations

This study has several limitations. First, the sample was small (22 providers and 130 coded interactions), so the clustered confidence intervals remain wide for some language comparisons. Second, the validation sample came from a hepatitis training program and half of the cases were hepatitis-related, which may limit external validity for broader primary-care populations. Third, we validated against human-coded checklists and transcript-based expert ratings rather than direct clinical observation. Fourth, a single coder produced the human coding that serves as that reference standard and no transcript was coded twice, so we cannot report the reliability of the standard itself. Fifth, one expert rated the Vietnamese transcripts and the other rated the English translations, so rater and language are perfectly confounded and the two expert columns cannot be read as a language contrast. Sixth, the fidelity audit measured each script deviation at the level of the interaction and not the individual rubric item. Seventh, text-only interaction omits non-verbal clinical cues. Eighth, although the scoring pipeline is deterministic and version-pinned, LLM behavior may still change across future model releases. Finally, the method requires internet connectivity and personal device access, which may limit deployment in the most resource-constrained settings.

### 4.2 Conclusion

This study demonstrates that LLM-driven conversational vignettes can produce a low-cost, flexible, and scalable method for measuring health care provider competencies. The method eliminates the need for in-person enumerators while preserving the open-ended interaction format that is the foundation of vignette validity. It is open-source, built on widely used survey software, and designed for adaptation to a wide variety of clinical contexts and languages. As LLM capabilities continue to improve, both the chatbot interactions and the automated data extraction are likely to become more reliable, further reducing the cost and increasing the feasibility of routine competency measurement. With further validation across settings and languages, this method has the potential to support rapid, low-cost measurement of health care provider competencies in diverse health system contexts.

## Acknowledgments

We acknowledge support in clinical vignette development, as well as transcript grading, from Dr. Marwatunnisa Al Mubarokah. We acknowledge additional support in vignette development and localization, as well as in leading the focus group and transcript grading, from Dr. Pham Thanh Thuy. We acknowledge the invaluable assistance, advice, and logistical coordination from Thu Huyen Nguyen and the rest of the staff at HAIVN. We acknowledge support in technical development of the survey plugin by Amrik Cooper and Mofya Phiri of Dobility, Inc. (SurveyCTO). We thank the participants in the focus group, management at Phu Tho Province General Hospital, and all respondents and StITCH program staff for their time, expertise, and feedback.

## Author contributions

**Conceptualization:** BD, DD. **Data curation:** BD, WSZ, HN. **Formal analysis:** BD. **Funding acquisition:** BD, DD. **Investigation:** BD, WSZ, HN. **Methodology:** BD, DD. **Project administration:** BD, HN, DD. **Resources:** DD. **Software:** BD. **Supervision:** BD, DD. **Validation:** BD, WSZ. **Visualization:** BD. **Writing – original draft:** BD. **Writing – review & editing:** BD, WSZ, HN, DD.

## Data availability statement

The de-identified data and the analysis code needed to reproduce every table and figure in this paper are deposited at Zenodo: https://doi.org/10.5281/zenodo.22103293. Interaction transcripts are not released. They are free-text clinical consultations recorded by a small number of identifiable providers in a single province, and the combination of case assignment, consultation content, and provider characteristics remains re-identifying after direct identifiers are removed. The chatbot fidelity audit reads those transcripts directly, so the numbers it reports in the Results cannot be recomputed from the deposited data; its code is deposited with the rest of the analysis.

## Competing interests

The authors declare no competing interests.

## Financial disclosure

This work was supported by the Harvard University Asia Center, the Harvard University Rose Service Learning Fellowship program, the Harvard University Institute for Quantitative Social Science, and Gilead Sciences. The funders had no role in study design, data collection and analysis, decision to publish, or preparation of the manuscript.

## Notes

### Competing Interest Statement

The authors have declared no competing interest.

### Author Declarations

This study was approved by the Ethical Review Board of the Hanoi University of Public Health (025-375/DD-YTCC) and the Mass General Brigham Institutional Review Board (Protocol 2023P003011). All participants provided written informed consent prior to participation.

### Summary of Updates

We revised the manuscript after peer review. We now report the automated transcript classification as item-level agreement and AUROC against human coding, using a speaker-attributed pipeline, with confidence intervals clustered on providers. We added an audit of chatbot fidelity across all fielded transcripts to the Results, and we report the focus group realism ratings with confidence intervals. The supplementary appendix was rebuilt. The de-identified data and analysis code are deposited at Zenodo, and author affiliations were updated.

