## Supplementary material for "Large language models for self-administered conversational vignette assessment of provider competencies: A pilot and validation study in Vietnam with automated LLM-powered transcript classification": S1 Appendix

#### Contents

**1 Clinical case scenarios and essential diagnostic checklists** **2**

**2 Supplementary tables** **41**

**3 Focus group supplementary results** **46**

**4 Automated scoring prompt** **49**

**5 Chatbot fidelity audit prompt** **50**

**6 STROBE checklist for cross-sectional studies** **52**

### 1 Clinical case scenarios and essential diagnostic checklists

The following pages contain the full clinical case scenario descriptions for all ten vignettes used in the study. Each case includes the patient background, chief complaint and history of present illness, medical and social history, the standardized opening statement, the full list of provider questions with pre-scripted standardized patient responses, clinical examination findings, laboratory test results, the preliminary diagnosis, recommended treatment, and counseling points. These scenarios were developed by expert clinicians on the research team and used as system prompts for the LLM-based chatbot to achieve standardized patient behavior across interactions.

#### Case 1: Asthma

**Patient Background**

Mai is 24 years old and is currently living with her aunt in a house on the edge of the city with 3 rooms. She moved into the city with her aunt a year ago when she finished university and has been looking for a job, but has not found one yet. Recently, she has been having a cough which worsens at night and in the early hours of the morning. Last night, Mai did not sleep well. Her cough seemed to worsen. She had bouts of difficulty breathing and her chest was producing a whistling sound. This seems to have been triggered by the rain and cold weather last night as she came home late, driving on a motorbike, and she was not dressed warmly.

Mai's parents are shopkeepers in the nearby province. All her family is in good health, except for her older brother who has had some breathing problems for the last couple of years and has been taking treatment for the same. She remembers her mother saying that her childhood was spent with episodes of coughing and breathing difficulties, but these episodes seemed to disappear in secondary school.

**Chief Complaint & History of Present Illness**

This problem started with an occasional episode a year ago, but over the last month, Mai has had breathing problems about once a week. This problem seems to get worse when there is dust in the air and cold weather. Earlier, the episodes used to last for a few minutes, but lately, they take about 10 – 15 minutes to get relief. The breathing problem is often accompanied by a dry cough, more so at night. During such episodes, she finds relief with a hot cup of ginger tea or warm water and sometimes takes cough syrup.

**Medical & Social History**

This morning, Mai was worried that the ginger tea did not work as fast as it usually does. She is exhausted. Her aunt is concerned about her condition and insists that Mai see a doctor for fear that her symptoms will worsen or recur at night.

**Opening statement:** Doctor, I have difficulty breathing that is worse this morning than it has been before.

**Provider Questions w/ SP Responses (case-specific)**

1. Does the difficulty breathing come and go and/or is it episodic?
   Yes.
2. How long does an episode or attack typically last?
   Most last 10-15 minutes but I do not feel completely well for an hour or so after.

3. Since when have you had these symptoms?
   It's been a couple of months, but since last month it has been getting worse.
4. Have you had any other episodes previously?
   Yes, it is now happening often since the season has changed.
5. Do you have a fever?
   No.
6. How often does it happen?
   Several times a week.
7. Do you cough?
   Yes, sometimes, but sometimes it is just hard to breathe.
8. Are you coughing a lot?
   No.
9. Tell me more about your cough. Is it dry or wet?
   It is dry.
10. Are you coughing up any blood or mucus?
    No, I am not coughing anything up.
11. Do you ever have wheezing/noise in your chest?
    Yes, there is a whistling noise, especially when it is bad..
12. Have you been in contact with anyone with fever and cough in the previous 3-5 days?
    No.
13. Have you lost weight?
    No.
14. Have you had night sweats?
    No.
15. Do you have any pain?
    No.
16. How do you get relief?
    Ginger tea has helped before but now it does not seem to work.
17. What triggers the episodes (e.g., dust, pollution, bad air quality, cold)?
    It happens when there is a lot of dust, but there is always a lot of dust here.
18. Does it happen when the weather is cold?
    Yes.
19. Does it happen when it is dusty?
    Yes.
20. Does it happen when it is smoky?
    Yes.
21. Does it change with the season change?
    I am not sure, I have only been living here one year.
22. Is it worse at certain times of the day?
    It is worse at night and sometimes it happens in the morning.
23. Do any of your siblings/parents have similar problems?

My brother has some breathing issues sometimes.

24. Does anyone in your household or family have a history of similar symptoms?

    My brother has some breathing issues sometimes.
25. Has this happened before when you were younger?

    Yes, but not for a long time so I don't know if it's the same thing.
26. Have you taken any medication?

    Yes, cough syrup.
27. Do you know the name of the cough syrup?

    No, I just got it from the pharmacy.
28. Do you smoke?

    No.
29. Do you drink alcohol?

    No.
30. Do you have hypertension / high blood pressure or diabetes/blood sugar issues?

    No.
31. Have you ever had any HIV/STIs/STDs? Any history of HIV/STIs/STDs?

    No.

**Clinical Examination Findings:**

1. Vital signs: Temperature is 36.2 C, blood pressure 114/74 mm Hg, pulse 65 beats per minute, and respiration 20 breaths per minute. Pulse oximeter: 95% in the room air.
2. Head and neck: lymph nodes not palpable.
3. Respiratory examination: both lungs are resonant by percussion. Auscultation reveals slight bilateral wheezing, no rhonchi.
4. Other examinations are normal.

**Tests:**

- Peak expiratory flow (PEF) : 71%
- Predicted FEV1: 79%

**Preliminary Diagnosis:** Asthma (mild persistent)

**Treatment Recommendations:**

- Treat as an outpatient
- Low-dose ICS-formoterol for symptom relief (e.g., Budesonide-fomoterol 160/4.5 mcd, 1 inhalation as needed)
- As-needed SABA

**Counseling point:**

- Explain possible triggers (i.e., dust, cold air), and the importance of limiting exposure to the triggers
- Use a mask, particularly in places or conditions where the trigger is present
- Keep the room clean and well-ventilated
- Explain conditions that required emergency management (e.g., worsening difficulty breathing)
- Explain how to use the medication (inhaler or spacer technique)

**Referral:** Not required unless condition worsens

#### Case 2: Pneumonia

**Patient Background:**

Minh is a 62 year old man. He previously had no significant health issues. He lives with his wife Anh, who is 60, and their youngest daughter, who is 25, in a two-bedroom apartment on the edge of the city. They also have another daughter and a son, who are 27 and 30, who live in the city. Minh works in an office in the city doing translation work in French and is looking forward to his retirement in the next few years. Anh worked as a schoolteacher until last year, when her son had a child and she retired, and she now cares for her son's 1-year-old child during most days. Minh's parents passed away several years ago in their home in the nearby province.

**Chief Complaint & History of Present Illness:**

Minh has been having a cough for five days, which has been consistent and is now causing him to be very tired. The cough has been productive with yellow mucus for the last three days. He also developed a fever and general malaise along with the cough and took the last two days off from his job, which is very unusual for him, and his wife is very worried. He has experienced chest pain, fever, and difficulty breathing, especially in the past two days. He could not sleep last night as he coughed continuously, and the right chest pain was worsening. He seems tired, haggard, and underweight. No family history of asthma. No family history of chronic cough.

**Medical & Social History:**

He took some tablets from the pharmacist. No known allergies. No previous history of chronic productive cough. Minh gave up smoking around 5 years ago. But previously, he had been smoking a minimum of 1 pack of cigarettes per day since he was 20.

**Opening Statement:** Doctor, I have been coughing for the last couple of days, and it has been worsening. For the past two days I took off work and my wife is worried.

**Provider Questions with SP Responses:**

1. What other symptoms do you have?

   Fever, chest pain, difficulty breathing.

2. How long has the cough lasted?

   Five days.

3. Is the cough dry or productive?

   Productive.

4. What color is the sputum/mucus?

Yellow.

5. Is there blood in the sputum/mucus?

   No

6. Is there chest pain?

   Yes.

7. Any difficulty in breathing?

   Yes.

8. How is your appetite?

   Decreased, I am very tired and don't want to eat much.

9. Do you have a high or low fever?

   It is mild, I am not having chills or sweating or anything.

10. Are you tired or lethargic?

    Yes.

11. Any difficulty in swallowing?

    No.

12. Is there a runny nose?

    No, but everything feels congested and it is hard to breathe.

13. Have you received any medication?

    Yes, I took some tablets from the pharmacist, just acetaminophen

14. Have you experienced similar symptoms before?

    Not that I can remember, I am a very healthy person

15. Do any of your siblings/parents have similar problems?
    No

16. Does anyone in your household or family have a history of similar symptoms?
    No

17. Redness of eyes?

No.

18. Have you received the COVID-19 vaccine?

    Yes

19. Have you received the influenza vaccine?

    No

20. Do you smoke?
    I gave up smoking 5 years ago. But I smoked around a pack of cigarettes per day starting when I was 20.

**Clinical Examination Findings:**

- Vital signs: Temperature is 39.2 C, blood pressure 154/80 mm Hg, pulse 105 beats per minute, and respiration 24 breaths per minute. Pulse oximeter: 93% in the room air.
- Head and neck: lymph nodes not palpable.
- Respiratory examination: both lungs are resonant by percussion, except the right lower lung fields are dull. Auscultation reveals bilateral diminished vesicular breath sounds, rhonchi, and late inspiratory crackles (are heard) in the area of the right lower lung fields. No wheezing.
- Other examinations are normal.

**Tests:**

- Chest X-ray: Patchy alveolar opacification in the right lower lobe.
- CBC: White blood cell, 17,000/mm3; neutrophils 70%, bands 15%, lymphocytes 15%, platelet: 200,000 µL, Hb: 12 g/dL

**Preliminary Diagnosis:** Pneumonia or community-acquired pneumonia

**Treatment Recommendations:**

- Treat as an inpatient.
- Ceftriaxone 2g daily, IV
- Paracetamol 500 mg PO up to four times daily
- Monitor his vital signs

**Counseling point:**

- Explain conditions that required emergency management (e.g., confusion, worsening difficulty breathing)
- Encourage feeding and fluid intake

**Referral:** Not required unless condition worsen

#### Case 3: Type II Diabetes Mellitus

**Patient Background:**

Tran, a 48-year-old man, has been obese for up to a decade. He works as a banker in a local bank and has been at this job for the last 25 years. He lives with his wife Loan and 2 sons in a three-bedroom apartment in the city. His wife is 50 years old and his sons are 21 and 23. His sons are both attending university part-time and they both also work driving motorcycles when they are not at school. Loan works as an office manager at a local office that coordinates research work with the government and with regional partners, so she travels often. His parents live in a town in another part of the country and are retired.

**Chief Complaint & History of Present Illness:**

Tran has had fatigue, frequent urination, increased thirst, and hunger for the past 3 months. He often wakes up at night due to the need to urinate. He also experienced occasional dizziness and mild breathlessness on exertion in the past 1 month. No prior medication.

**Medical & Social History:**

No smoking or exercise. No previous history of chronic illness. No prior treatment or medications. The past HIV test was negative. He occasionally drinks alcohol, once a week or once in two weeks. He almost eats fast food every day for his lunch because of his short break during the day. He is 165 cm tall and 82 kg body weight.

**Opening Statement:** Doctor, I feel weak recently going about my day. I have been thirsty and hungry more often than usual and this is concerning my wife.

**Provider Questions with SP Responses:**

1. How long has this been going on?

   I have been feeling especially tired and thirsty over the last month or so.

2. Do you have a fever?

   No.

3. Do you have any vomiting?

   No.

4. Has your appetite changed?

   Yes, it increased.

5. Has your thirst changed?

   Yes, it increased.

6. Do you have diarrhea?

   No.

7. Do you have a cough?

   No.

8. Do you experience shortness of breath?

   Yes, when climbing stairs.

9. Do you take any medications?

   None.

10. How about your urination?

    It has become more frequent. I also often wake up at night with the need to use the toilet.

11. Do you experience numbness or tingling in your limbs?

    No.

12. Do you smoke?

    No.

13. Do you consume alcohol?

    I occasionally drink alcohol, about once a weekend I will go out with friends or with my sons to watch sports and we will have 2-3 beers throughout the afternoon.

14. Do you regularly exercise?

    No.

15. Did you take any health checks?

    Yes, about three months ago I had a routine screening and they advised me to be aware of blood sugar and blood cholesterol but I have not done anything more with the doctors.

16. How about blood pressure? Do you have a history of high blood pressure?

    I don't think so.

17. Do you have any family history of diabetes?

    I don't think so.

18. Do you have any family history of hypertension?

    I don't think so.

19. Do you experience any dizziness?

    Yes, sometimes.

20. Do you have any headaches or joint pain?

    No

**Clinical Examination Findings:**

- Vital signs: Temperature is 36.5°C, blood pressure 124/83 mm Hg, pulse 75 beats per minute, and respiration 14 breaths per minute
- Head and neck: lymph nodes not palpable
- Abdomen: Normal
- Oral exam: Normal
- Neurological exam: Normal
- Fundoscopy: No changes
- Weight: 82kg
- Height: 165cm

**Tests:**

- Fasting blood sugar: 12.8 mmol/L
- Random blood sugar: 16.7 mmol/L
- HbA1c: 6.9

**Preliminary Diagnosis:** Type II Diabetes Mellitus

**Treatment Recommendations:**

1. Refer to the diabetic clinic
2. Start oral hypoglycemics
3. Lifestyle modifications: diet and exercise

4. Monitor glucose levels

**Counseling points:**

1. Explaining his condition and diagnosis
2. Explain the importance of foot care and how to do that
3. Instructions about diet and exercise guidance
4. Medication adherence

**Referral:** Yes, for diabetic management

#### Case 4: Pulmonary Tuberculosis

**Patient Background:**

Toan is a 40-year-old manual laborer. He works with concrete and rebar at various small construction jobs on houses, sidewalks, and other residential and commercial projects. He lives with his wife My, who is 35, and his elderly father, who is 70, in a small house with very little ventilation at the edge of the city in a crowded district. His mother passed away some years ago. Toan and My have no children and have been married since he was 25 years old. Toan has worked and lived here since he finished school at 18 years old.

**Chief Complaint & History of Present Illness:**

Toan has had a persistent, productive cough, with occasionally a bit of blood in the sputum for the past month. The cough occurs persistently and does not change in a particular condition (i.e, night, cold, dusty). He has also experienced low-grade fever, night sweats, and fatigue on and off. He thinks that he might be losing weight as his work trousers feel looser, but it has been hot and he has also not been very hungry recently. He has not had a chronic illness before. No prior medication. No previous HIV testing.

**Medical & Social History:**

He smokes a pack of cigarettes daily, although this has decreased since he started coughing. Family history of chronic cough (father). Alcohol use ceased two years ago.

**Opening Statement:** Doctor, I've had a cough for weeks now and it won't stop. I constantly feel tired.

**Provider Questions with SP Responses:**

1. Since when have you had this cough?

   About one month.

2. What is the cough type?

   Productive with sputum.

3. What is the color of the sputum?

   Usually yellowish

4. Is there any blood in the sputum?

   Yes, every now and then.

5. What is the color of the blood?

Light red

6. Is the cough a change in a particular condition, such as when it is cold?

   No, it's persistent throughout the day.

7. Do you experience chest pain or breathing difficulty?

   No

8. Do you have a fever? If yes, is it high or low?

   Yes, low fever sometimes.

9. How was the fever pattern?

   Low-grade (not really severe), and sometimes it is worse at night.

10. Do you have any night sweats?

    Yes.

11. Does anyone in your household have a chronic cough?

    My father has had a prolonged cough for many years.

12. Has your father gotten treatment?

    No

13. Do you experience weight loss?

    I think so, but it has been hot and I have not really been hungry.

14. Does your appetite change?

    Yes, it decreased.

15. Do you have fatigue?

    Yes.

16. Have you ever had a similar cough like this before (prior episodes)?

    No.

17. Do you have a history of previous medication?

No.

18. How about your alcohol use?

    I quit 2 years ago.

19. Do you smoke?

    Yes, about one pack/day.

20. Have you ever tested for HIV?

    No

21. How about your diet?

    I usually eat rice, vegetables, and occasional meat.

22. What is your profession?

    Manual laborer.

23. Have you had sex with a sex worker previously? Any high-risk sexual behavior?

    Yes, but not since I was married.

24. Have you ever engaged with IV drug use?

    No

- **Clinical Examination Findings:**
- Patient alert
- Vital signs: Temperature: 38°C Pulse: 90 bpm, Respiratory rate: 20x/minutes, Blood pressure: 120/80
- Weight: 53 kg Height: 165 cm
- Head and neck: no lymphadenopathy
- Respiratory examination: Chest auscultation: crackles in the upper lobe of the left lung

**Tests:**

- Sputum exam: Positive for AFB
- Chest X-ray: Hilar lymphadenopathy, consolidation, cavities
- ESR: High Hemoglobin: 9 g/L
- HIV test: Negative
- Blood sugar test: Normal

**Preliminary Diagnosis:** Pulmonary Tuberculosis

**Treatment Recommendations:**

- Start TB regimen: 4 drugs for 2 months, then 2 drugs for 6 months
- Refer to the TB clinic for follow-up

**Counseling points:**

- Adherence to TB medication
- Testing and treatment for close contacts
- Hygiene and ventilation
- Nutrition guidance

**Referral:** Yes, for full TB and possible HIV testing and management

---

#### Case 5: Hypertension

**Patient Background:**

Dang, 54 years old, is an office manager in a small factory that makes sports shoes and accessories. He has been having frequent headaches for the past 3 months. He did not have a significant health issue previously. He lives with his wife Linh, who is 50, and their one son, who is 25, in a two-bedroom apartment. Their son is in school. His parents live nearby and are retired from similar jobs, and Dang looks after them often.

**Chief Complaint & History of Present Illness:**

Dang reports headaches for the past 3 months, mainly in the mornings, described as dull and persistent. The headaches become worse, especially when he is under stress at work or when he lacks sleep due to caring for his parents. He reports that the headaches have been getting more frequent for the past 3 weeks. He denies visual changes, chest pain, palpitations, or shortness of breath. He occasionally feels dizzy and tired in the late afternoon. He is not aware of any triggers. The headache sometimes occurs with nausea, vomiting, or heartburn.

**Medical & Social History:**

No prior treatment or medications. No known allergies. No previous history of chronic disease. He often consumes fast food because of frequent overtime at work. Non-smoker, and drink beer usually on weekends. Very rare to do physical exercise. Father has hypertension that is managed with medication, and mother has type 2 diabetes.

**Opening Statement:** Doctor, I have been having a headache on and off for a while now.

**Provider Questions with SP Responses:**

1. What other symptoms do you have?

    I also often get tired in the afternoon and take a nap after lunch. Sometimes I also have dizziness, nausea, or heartburn, but it varies.

2. Can you describe how the headache is?

    It is dull and persistent.

3. When does it usually start during the day?

    In the morning when I wake up for work.

4. How many days has it been?

On and off for 3 months but getting worse recently.

5. Does the headache come every day?

    Almost every day, especially for the past 3 weeks

6. Is there chest pain?

    No but sometimes there is heartburn.

7. Any difficulty in breathing?

    No

8. Any visual changes?

    No

9. Any heart palpitations?

    No

10. Do you have numbness or tingling in the limbs?

    No

11. Do you feel tired easily?

    Yes, I need a nap almost every afternoon.

12. Do you have a previous disease or illness?

    None

13. Do you have a history of diabetes?

    No

14. Do you have a history of high blood pressure?

    No

15. Do you have a history of stroke?

    No

16. Do you have any family history of hypertension?

My father takes medication for this.

17. Do you regularly exercise?

    No

18. How about your everyday food?

    I often eat street food at work and on my way home if I work late.

19. Have you received any medication?

    No

20. Did you take any health checks before?

    No

**Clinical Examination Findings:**

- Vital signs: temperature is 36.8°C, blood pressure 158/96 mm Hg (repeated twice), pulse 84 beats per minute, and respiration 16 breaths per minute
- Weight: 86 kg, height: 170 cm. BMI 29.8 (overweight)
- The other physical examination is normal

**Laboratory Test:**

- Haemoglobin (Hb): 151 g/L, Hct = 41%
- Lipid profile: total cholesterol: 13.6 mmol/L, LDL: 7.88 mmol/L, HDL: 2.1 mmol/L, triglycerides: 7.77 mmol/L
- Renal function: creatinine: 97.26 µmol/L, sodium (Na): 142 mmol/L, potassium (K): 4.1 mmol/L, Chloride (Cl): 103 mmol/L
- ECG: sinus rhythm, HR 84 bpm, no ischemic changes, mild left ventricular hypertrophy
- Urinalysis: protein (-), glucose (-), pH 6.0, RBC/WBC normal

**Preliminary Diagnosis:** Stage 1 hypertension, dyslipidemia

**Treatment Recommendations:**

- Treat as an outpatient.
- Give Antihypertension (ACEI inhibitor or CCB) for 2 weeks.
- Initiate statin treatment
- Recheck BP in 1 month.

**Counseling point:**

- Instruct on how to drink the medicine
- Lifestyle modification (DASH diet-low sodium, high fruits, low saturated fat, exercise regularly, reduce salt intake, and alcohol)
- Instruct to return after 2 weeks
- Explain conditions that needs emergency care

**Referral:** Not required unless condition worsens

#### Case 6: Hepatitis B with Cirrhosis

**Patient Background:**

Tuan, a 64-year-old retired farmer, was referred to your facility with fatigue and yellowing of the eyes. His symptoms have been progressing for a month, including loss of appetite and a 2 kg weight loss. He occasionally consumes alcohol. He has never smoked and lives with his wife in a rural area, and has come to your clinic after seeing a health worker. His son, who is 35, lives in the city and works in construction.

**Chief Complaint & History of Present Illness**

The health worker ordered a blood test for him through a local clinic and he got an HBsAg+ result from the testing, but does not know what this means for his health. Tuan's condition has been progressing for 1 month; he has fatigue, loss of appetite, lost 2 kg/month, accompanied by yellow eyes, gradually increasing jaundice, dark yellow urine, swelling of the leg, and the belly getting bigger. The patient has no fever, vomiting, bleeding, or difficulty breathing, has not received any treatment at home, and has now come to your facility after being referred by the primary care clinic via the health worker in his area.

**Medical and Social History**

He never complained of any significant jaundice or fatigue to his wife before. The condition has been progressing for around 1 month; the patient has fatigue, loss of appetite, weight loss of around 2 kg, no fever, and has not taken any medication at home. The patient has occasionally drunk alcohol for the past 20 years, 3 glasses of wine each time, and does not smoke. He does not know his family history of hepatitis or liver disease.

**Opening statement:** Doctor, I have been having fatigue, loss of appetite, and yellowing eyes for a while. I was referred by my usual health worker because of the result from a lab test they asked me to do.

**Provider Questions with SP Responses:**

1. Have you ever been diagnosed with hepatitis B?
   No. But my laboratory test was HBsAg+ from my primary care provider
2. Have you ever taken any antiviral medications?
   No.
3. Do you consume alcohol?
   Occasionally if we have dinner with friends, about 3 glasses of wine each time
4. Do you smoke?
   No.
5. Have you had any recent weight changes?
   Yes, I lost about 2 kg
6. Has your appetite changed?
   Yes, it decreased

7. Have you noticed any changes in your skin?
   Yes, my eyes and skin turned yellow
8. How about your urine color?
   My urine also became dark yellow
9. Since when have you experienced these symptoms?
   It has been around one month
10. Do you experience any vomiting?
    No
11. Do you experience any bleeding?
    No
12. Do you experience any difficulty breathing?
    No
13. Do you have a history of chronic illnesses or liver diseases?
    No
14. Do you have a fever?
    No
15. Do you have leg swelling?
    Yes
16. Do you think that your belly is getting bigger?
    Yes
17. Do you have a history of sexually transmitted diseases (STDs)?
    No
18. Do you have a history of using unsafe injections?
    No
19. Any history of liver disease in your family?
    I do not know.
20. How about your wife or child? Do they have any hepatitis B or C?
    I don't think so
21. Any medications at home currently?
    No.

**Examination Findings:**

- Patient is alert
- Vital signs: temperature is 97.4°F, blood pressure 118/76 mm Hg, pulse 64 beats per minute, and respiration 15 breaths per minute
- Height: 175 cm, weight: 60 kg
- Head and neck: Yellow eyes, yellow skin, no lymphadenopathy
- Extremities: bruises on hands, leg edema
- Abdominal examination: Distended abdomen with dullness to percussion in the lower area (ascites), Palpable spleen (2.5 cm below the costal margin), no palpable liver
- Normal other exams

**Laboratory & Imaging findings:**

- Total bilirubin: 65.2 µmol/L, Direct bilirubin 32.5 µmol/L, Albumin: 28 g/L
- AST/ALT: 78/64 IU/L, INR: 1.78
- HBV DNA: 2490 IU/mL, HBeAg negative, anti-HBe positive
- Anti-HCV negative
- HIV negative
- Platelets: 98 x 10^9/L
- AFP: 12.8 ng/mL, Creatinine: 77 µmol/L
- Ultrasound: Fibrosis, splenomegaly, ascites, no nodules in the liver
- Gastroscopy: Grade 3 esophageal varices

**Preliminary diagnosis:** Chronic hepatitis B virus with decompensated cirrhosis (Child-Pugh Class C).

**Treatment Recommendation:**

- Hospital admission
- Initiate TDF 300 mg/day (CrCl 72.79)
- Monitor for side effects (renal, lactic acidosis, osteoporosis)
- Improve cirrhosis status and prolong life expectancy
- Varices ligation
- Propranolol 40 mg PO daily

**Monitoring Plan:**

- Hospitalize until ascites and jaundice resolve
- Clinical and liver function monitoring every 2 weeks
- HBV DNA every 6 months
- Viral load and Anti-HBe at week 12
- Liver cancer/HCC screening every 12-24 weeks
- Fibrosis evaluation every 12-24 weeks
- Creatinine monitoring every 6 months

**Counseling Points:**

- Explain hepatitis B, the risk of progression to liver cancer, and bleeding from esophageal varices and other complications
- Importance of lifelong treatment, adherence, and follow-up
- Screening and vaccination for family members
- Lifestyle changes: stop alcohol, healthy diet, rest, exercise

#### Case 7: Hepatitis B in Pregnancy

**Patient Background:**

Pham is a 31-year-old schoolteacher, currently 24 weeks pregnant with her second child. They do not know the child's gender. Pham was diagnosed with hepatitis B several years ago, but has not received any treatment and wants to make sure she is okay at this stage in her pregnancy. She has tested HBsAg+ for several years. Her husband is 35 years old and works in a nearby office for a transportation and tourism service. They have a 2-year old girl Hue currently who is healthy and developing normally. Pham has not had any apparent liver symptoms or complications from her condition, and her pregnancy is otherwise normal. This is her second pregnancy. Her husband and daughter have never been tested for hepatitis.

**Chief Complaint & History of Present Illness**
This is her second pregnancy, and the first child never got tested for Hepatitis B. Pham has no underlying diseases and is not taking any medications. Pham does not experience fatigue, loss of appetite, yellow eyes, or any bleeding. No liver enzymes and virus count tests were done, so no medications were taken. HIV and syphilis tests are negative.

**Medical & Social History**
No one has cirrhosis or HCC in Pham's family. Her husband has never been tested with Hepatitis B or other liver-related illnesses.

**Opening statement:** Doctor, I was diagnosed with hepatitis B a few years ago, and now that I am pregnant with my second child, I wanted to come here and check to make sure everything is okay.

**Provider Questions with SP Responses:**

1. When did you find out about hepatitis B?

   A few years ago.

2. How do you know about your hepatitis B status?

   My HBsAg was positive a few years ago

3. Besides HBsAg, are there other laboratory tests, such as liver enzymes or virus count, that you took before?

   I don't think so

4. Have you received any antiviral treatment?

   No.

5. Do you experience any symptoms?

   No.

6. Do you have any fatigue?

   No

7. Do you have a loss of appetite?

   No

8. Have you ever had yellow eyes or skin?

   No

9. Do you have any bleeding?

   No

10. Do you have other health conditions or medications?

    No.

11. How about your current pregnancy condition?

    I regularly get antenatal care (ANC), and the result is normal

12. Is there anyone in your family who has liver disease?

    I don't think so

13. Has your family been tested for hepatitis before?

    I don't think so

14. How about the hepatitis B status of your husband and children?

    They have not yet been tested

15. In your first pregnancy, did you get any viral tests or other health checks?

    No

16. Do you know your HIV status?

    It is negative

17. Have you taken a syphilis test?
    It is negative
18. Do you consume alcohol?
    No
19. Do you smoke?
    No

**Examination findings:**

- The patient is alert
- Vital signs: stable
- Skin and extremities: No jaundice, bruising, or edema, no subcutaneous hemorrhage
- Abdominal examination: Normal liver/spleen palpation
- Other examination: normal
- Fetal growth is appropriate for gestational age

**Laboratory & Imaging findings:**

- PLT: 250 G/L, HGB: 130 g/L
- AST/ALT: 19/20 U/L
- HBeAg: Negative, HBV DNA: 200 IU/mL
- Anti-HCV and HIV: Negative
- Ultrasound: Normal liver, spleen, and fetus is following gestational age

**Preliminary Diagnosis:** Chronic hepatitis B without fibrosis

**Interpretation:**

- APRI score = 0.3 → no fibrosis
- HBeAg negative and HBV DNA: 200 IU/mL → very low viral activity
- No symptoms, normal imaging

**Treatment Recommendation:**

- Manage in outpatient care
- No antiviral needed (not meeting treatment threshold)
- No prophylaxis needed (HBV DNA < 200,000 IU/mL)

**Monitoring Plan:**

- Mother: re-evaluate 3 months after delivery or earlier if symptoms occur
- Baby: HBV vaccine and HBIG within 24 hours after birth; 3 HBV doses in Expanded Program on Immunization (EPI); HBsAg and anti-HBs from month 7

**Counseling Points:**

- Explain the current disease status and the need for monitoring
- Infant protection protocol
- Encourage family testing
- Breastfeeding is safe
- Choose to disclose status if desired

#### Case 8: HBV Not Eligible for Treatment

**Patient Background:**

Tran is a 44-year-old store manager who was diagnosed with chronic hepatitis B 29 years ago. He's asymptomatic and attends biannual check-ups. He was diagnosed at age 15. He lives with his wife, who is 39, and 2 of their 3 sons in a three-bedroom apartment. They recently moved to this area when he got a promotion and his two children who live with him have started at a local school. His oldest has just gone to university.

**Chief Complaint & History of Present Illness**

Tran is currently very healthy and has never experienced any significant liver-related symptoms. He has been diagnosed with chronic Hepatitis B infection for 29 years and has never been on antiviral treatment. He just transferred to your facility because of the family relocation, and he visited for the first time for a regular check-up for his hepatitis B, which is scheduled to take place every 6 months. Family history: mother had hepatitis B.

**Medical & Social History**
He did not use injection drugs, smoked, and only drinks alcohol 1-2 times/month with 2-3 glasses of beer each time. He was previously tested negative for HCV and HIV. His mother was diagnosed with Hepatitis B. His siblings have never been tested with Hepatitis B. He was already married and had 3 children, and they had never been tested for Hepatitis B.

**Opening statement:** Doctor, I have had chronic hepatitis B for many years and I recently moved to this area, so I am going to be seeing you from now on. It has been about 6 months since my last exam.

**Provider Questions with SP Responses:**

1. When did you find out about hepatitis B?

   29 years ago.

2. How do you know about your hepatitis B status?

   I was diagnosed when I was 15 years old

3. Besides HBsAg, are there other laboratory tests that you took before?

   Yes, for my routine check-up

4. Have you received any antiviral treatment?

   No, because my previous doctor always said that I am not eligible for antiviral medication

5. Do you currently experience any symptoms?

   No.

6. Do you have any fatigue?

   No

7. Do you have a loss of appetite?

   No

8. Do you have yellow eyes or skin?

   No

9. Do you have any bleeding?

   No

10. Do you have other health conditions?

    No.

11. Is there anyone in your family who has liver disease?

    My mother was diagnosed with hepatitis B

12. Has your family been tested for hepatitis before?

    My mother

13. Has your mother or anyone else in your family been diagnosed with HCC?

    No

14. How about the hepatitis B status of your wife and children?

    They have not yet been tested

15. Do you know your HIV status?

    It is negative

16. Do you know your HCV status?

    Negative

17. Are you taking any medications?

    No.

18. Do you smoke or drink?

    Occasionally drinks beer, usually 2-3 glasses each time, no smoking.

19. Do you ever use any injection drugs?

    No

20. Do you have any history of sexually transmitted diseases (STDs)?

    No

21. Do you have a history of any risky behaviour?

    No

**Examination Findings:**

- The patient is alert
- Vital signs: stable vitals
- Skin and extremities: Normal skin and mucous membrane, no jaundice or edema
- Abdominal examination: Normal liver/spleen
- Other examination normal

**Laboratory & Imaging findings:**

- PLT: 280 G/L, HGB: 140 g/L
- AST/ALT: 20/28 IU/L
- HBeAg: Negative, HBV DNA: 30 IU/mL
- Abdominal Ultrasound: liver normal, no nodules or fibrosis

**Preliminary Diagnosis:** Chronic hepatitis B without fibrosis

**Treatment Recommendation:**

- Not eligible for treatment
- No fibrosis or significant viral activity
- No additional tests of liver function are necessary

**Monitoring Plan:**

- Follow up every 6-12 months

- Clinical exam, LFTs, fibrosis, HBV DNA, and HBeAg

**Counseling Points:**

- Disease is stable, no treatment needed
- Importance of regular monitoring
- Screen family
- Avoid alcohol and live a healthy lifestyle

#### Case 9: Hepatitis C

**Patient Background:**

Ngo is a 28-year-old auto mechanic who discovered he was anti-HCV positive after a blood donation he made two years ago. He never followed up until recently, because he began experiencing fatigue, joint pain, and nausea for the past 3 weeks. He is unmarried and lives in a small house by himself. He has lived here since he finished school at 19 years old. He had a blood transfusion after a traffic accident when he was 15.

**Chief Complaint & History of Present Illness**
He experienced a notable loss of appetite and nausea for the past 3 weeks, which has made it hard to do his job. Besides that, he also often feels pain in his knee joints and weakness in his legs. However, he had no fever, no shortness of breath, and no jaundice or yellow eyes. As he felt uncomfortable, he sought traditional medicine, but his condition did not improve. Then he went to the clinic, and previous tests revealed that liver enzymes were also elevated, so he went to your facility.

**Medical & Social History**
He received a blood transfusion of red blood cells because of his accident 15 years ago. He hasn't been vaccinated against Hepatitis B, hasn't taken any drugs and does not drink alcohol, and does not smoke. Family medical history: no HCC and Hepatitis B, and C.

**Opening statement:** Doctor, I recently have felt very tired. My knees hurt, and I have had nausea. I went to a small clinic before coming here where they did some tests but my problems have not been resolved.

**Provider Questions with SP Responses:**

1. How long have you been experiencing these symptoms?

   For the past 3 weeks.

2. What symptoms have you noticed?

   Loss of appetite, nausea, joint pain (particularly knee joints), fatigue, weakness in my leg

3. Do you have any fever or shortness of breath?

   No

4. Do your skin or eyes become yellow?

   No

5. How about your urine color?

It is normal

6. Do you have leg swelling?

   No

7. Do you have a fever?

   No

8. Do you have any history of chronic illnesses or medications?

   No

9. Any history of blood transfusions or surgeries?

   Yes, a transfusion after an accident 13 years ago

10. Have you used traditional or herbal medicine for your current symptoms?

    Yes, but it didn’t improve.

11. Have your family members been tested with Hepatitis?

    No.

12. Do you have any past history of liver disease?

    I got my anti-HCV test positive during blood donation 2 years ago

13. Did you get antiviral treatment?

    No

14. For your current condition, have you taken any previous laboratory tests?

    Yes, from the clinic before I came here, the liver enzyme test results were elevated

15. Have you been vaccinated against Hepatitis B?

    No

16. Do you have any family members who have liver disease?

    No

17. Do you drink alcohol or smoke?

No

18. Do you know your hepatitis B status?

    No

19. Do you know your HIV status?

    No

20. Do you have a history of using unsafe injections?

    No

**Examination findings:**

- Patient is alert
- Vital signs: temperature is 97.7°F, blood pressure 114/73 mm Hg, pulse 71 beats per minute, and respiration 14 breaths per minute
- Skin and extremities: Mild yellowing of eyes, purpura on both legs, no jaundice, edema
- Abdominal examination: Normal

**Laboratory & Imaging findings:**

- PLT: 120 G/L, HGB: 125 g/L
- AST/ALT/GGT: 64/57/70
- Bilirubin: 30, Albumin: 32
- Anti-HCV: Positive, HCV RNA: 3.6 million IU/mL
- HIV, HBsAg: Negative, anti HBs 200 IU/mL
- Ultrasound: Normal

**Preliminary Diagnosis:** Chronic HCV with advanced fibrosis (F3), without cirrhosis and without co-infections

**Treatment recommendation:**

- Eligible for DAA therapy: SOF/VEL or SOF+DAC for 12 weeks
- Outpatient treatment

**Monitoring Plan:**

- After 1 month: LFTs, CBC
- SVR12: HCV RNA, fibrosis, AFP, ultrasound, LFT, CBC
- Screen for HCC every 6 months after SVR 12 evaluation (as he has F3 fibrosis)

**Counseling Points:**

- Explain the disease, purpose, and benefits of treatment
- Importance of adherence and side effect awareness
- Prevent reinfection
- Screen spouse and child
- Importance of seeking health services earlier and awareness of utilizing unstandardized traditional medicine

#### Case 10: Hepatitis C with ARV

**Patient Background:**

Dang is a 30-year-old living with HIV, and has been on antiretroviral therapy (ARV) since turning 21. He tested positive for HBsAg and anti-HCV at the time of HIV diagnosis but was never treated for hepatitis C. He works on a farm in the province, and is married to his wife Huong (30) for the last 6 years, and they have 3 young children, aged 2, 3, and 5. He now presents with jaundice and fatigue. He was an IV drug user who stopped 7 years ago, right before getting married, a little after he was diagnosed with HIV and started on ARV. He has no family history of liver disease. He is currently on TLD ARV regimen, with good antiretroviral therapy adherence. He has had an undetectable viral load and his wife and children have never been diagnosed with HIV.

**Chief Complaint & History of Present Illness**
He is a regular patient in your health facility. He has had yellowing eyes for the last 2 months, has also been tired almost all day. He has no fever, but often has nausea and sometimes vomiting. He has normal bowel movements and decreased appetite. There was no family history of liver cancer or Hepatitis B or C among his mothers, siblings, wife, and children.

**Medical & Social History**
He was diagnosed with HIV in November 2017 and is on ARV treatment, a TDF/3TC/EFV regimen. When he was enrolled, he was tested and got anti-HCV (+) and HBsAg (+). At that time, he was not offered an HCV RNA test or antiviral treatment. He has a history of using IV drugs when he was young, but has stopped for the past 7 years.

**Opening statement:** Doctor, I feel tired all the time, and my eyes have been a bit yellow since this happened. I am on antiretroviral therapy, is this related?

**Provider Questions with SP Responses:**

1. Have you had any symptoms recently?

   Yes, yellow eyes, nausea, vomiting, and tiredness.

2. Since when have you experienced those symptoms?

   2 months

3. Do you experience a fever?

   No

4. Do you experience leg edema?

No

5. Do you experience bleeding?

   No

6. How about your urine color?

   I think it is as usual

7. How about your appetite?

   It decreased

8. Do you have diarrhea?

   No

9. Do you have a chronic productive cough?

   No

10. Since when have you been taking ARV treatment?

    Since I turned 21.

11. What regimen that you take?

    TDF/3TC/EFV

12. Are you taking your HIV treatment regularly?

    Yes, daily.

13. Do you have a history of liver disease?

    When I enrolled in ARV treatment, I was tested and got anti-HCV (+) and HBsAg (+)

14. Besides that, did you take other laboratory tests such as an HCV RNA test?

    No

15. Did you take any antiviral or hepatitis medication?

    No

16. Do you drink alcohol?

No.

17. Do you have a history of using injection drugs?

    Yes, when I was young. But I have stopped for 7 years, since I got married.

18. Do you have any family members with a history of liver cancer or Hepatitis B, or C?

    No

19. Have your family members been tested for hepatitis?

    No.

**Examination findings:**

- Patient is alert
- Vital signs: temperature is 96.7°F, blood pressure 117/76 mm Hg, pulse 75 beats per minute, and respiration 15 breaths per minute
- Skin and extremities: mild jaundice, no edema, no subcutaneous hemorrhage
- Head and neck: No lymphadenopathy, no signs of fungal infection
- Abdominal examination: Abdomen is soft, liver and spleen are not palpable, no ascites or organomegaly
- Other examinations are normal

**Laboratory & Imaging findings:**

- PLT: 126 HGB: 120, AST/ALT: 100/70, GGT: 55
- Total bilirubin: 38, Albumin: 31
- HCV RNA: 2.1 million IU/mL
- HBsAg Positive
- HBV DNA: Not detected
- CD4: 400
- Creatinine: 80 µmol/L
- HIV RNA: not detected
- Ultrasound: Normal

**Preliminary Diagnosis:** Chronic viremic HCV infection, co-infected with HBV and HIV, compensated liver cirrhosis

**Treatment Plan:**

- Continue ART (switch from TDF/3TC/EFV to TDF/3TC/DTG (TLD) regimen)
- Begin SOF + DAC x 24 weeks

**Monitoring Plan:**

- 4-week follow-up: LFTs, CBC, creatinine
- SVR12 testing after 12 weeks post-treatment
- Continue HCC screening every 6 months

**Counseling Points:**

- Explain disease status and treatment benefits
- Importance of adherence to ARV and DAA
- Monitor for interactions and side effects
- Encourage partner and family testing
- Emphasize a healthy lifestyle and prevention of reinfection

#### 2 Supplementary tables

**Table S1. Case scenarios and focus group realism ratings.** Each row shows the case condition, the standardized opening statement the simulated patient delivered, and the recommended treatment and counseling points written into the case script, followed by mean focus group ratings on a 1–5 scale for overall realism, realism given the diagnosis, realism of the patient's social background, and case easiness. *n* is the number of non-missing realism ratings for that case.

| Condition | Opening statement | Recommended treatment and counseling | Overall | Diagnosis | Social | Ease | *n* |
|---|---|---|---|---|---|---|---|
| Asthma | Doctor, I have difficulty breathing that is worse this morning than it has been before. | Treat as an outpatient; low-dose ICS-formoterol for symptom relief (e.g., Budesonide-fomoterol 160/4.5 mcd, 1 inhalation as needed); as-needed SABA; explain possible triggers (i.e., dust, cold air), and the importance of limiting exposure to the triggers; recommend using a mask, particularly in places or conditions where the trigger is present, recommend keeping the room clean and well-ventilated, wxplain conditions that required emergency management (e.g., worsening difficulty breathing); explain how to use the medication (inhaler or spacer technique) | 3.86 | 3.86 | 3.43 | 3.71 | 7 |
| Pneumonia | Doctor, I have been coughing for the last couple of days, and it has been worsening. For the past two days I took off work and my wife is worried. | Treat as an inpatient; Ceftriaxone 2g daily, IV ; Paracetamol 500 mg PO up to four times daily; monitor his vital signs; explain conditions that required emergency management (e.g., confusion, worsening difficulty breathing); encourage feeding and fluid intake | 3.75 | 3.88 | 3.38 | 3.75 | 8 |
| Type II Diabetes Mellitus | Doctor, I feel weak recently going about my day. I have been thirsty and hungry more often than usual and this is concerning my wife. | Refer to the diabetic clinic; start oral hypoglycemics; recommed ifestyle modifications: diet and exercise; monitor glucose levels; explaining his condition and diagnosis; explain the importance of foot care and how to do that; instructions about diet and exercise guidance; emphasize medication adherence | 3.88 | 3.88 | 3.75 | 3.88 | 8 |
| Pulmonary Tuberculosis | Doctor, I've had a cough for weeks now and it won't stop. I constantly feel tired. | Start TB regimen: 4 drugs for 2 months, then 2 drugs for 6 months; refer to the TB clinic for follow-up; emphasize adherence to TB medication; recommend testing and treatment for close contacts; recommend hygiene maintenance and ventilation; provide utrition guidance | 3.67 | 3.78 | 3.78 | 3.67 | 9 |
| Hypertension | Doctor, I have been having a headache on and off for a while now. | Treat as an outpatient; give Antihypertension (ACEI inhibitor or CCB) for 2 weeks; initiate statin treatment; recheck BP in 1 month; instruct on how to drink the medicine; lifestyle modification (DASH diet-low sodium, high fruits, low saturated fat, exercise regularly, reduce salt intake, and alcohol); instruct to return after 2 weeks; explain conditions that needs emergency care | 3.75 | 3.88 | 3.88 | 3.62 | 8 |
| Hepatitis B with Cirrhosis | Doctor, I have been having fatigue, loss of appetite, and yellowing eyes for a while. I was referred to by my usual health worker because of the result from a lab test they asked me to do. | Hospital admission; initiate TDF 300 mg/day (CrCl 72.79); monitor for side effects (renal, lactic acidosis, osteoporosis); improve cirrhosis status and prolong life expectancy; varices ligation; propranolol 40 mg PO daily; hospitalize until ascites and jaundice resolve; clinical and liver function monitoring every 2 weeks; HBV DNA every 6 months; viral load and Anti-HBe at week 12; liver cancer/HCC screening every 12-24 weeks; fibrosis evaluation every 12-24 weeks; creatinine monitoring every 6 months; explain hepatitis B, the risk of progression to liver cancer, and bleeding from esophageal varices and other complications; explain importance of lifelong treatment, adherence, and follow-up; screening and vaccination for family members; explain lifestyle changes: stop alcohol, healthy diet, rest, exercise | 4.00 | 4.00 | 4.00 | 4.00 | 2 |
| Hepatitis B in Pregnancy | Doctor, I was diagnosed with hepatitis B a few years ago, and now that I am pregnant with my second child, I wanted to come here and check to make sure everything is okay. | Manage in outpatient care; no antiviral needed (not meeting treatment threshold); no prophylaxis needed (HBV DNA < 200,000 IU/mL); monitor other: re-evaluate 3 months after delivery or earlier if symptoms occur; monitor baby: HBV vaccine and HBIG within 24 hours after birth, 3 HBV doses in Expanded Program on Immunization (EPI), HBsAg and anti-HBs from month 7; explain the current disease status and the need for monitoring; explain infant protection protocol; encourage family testing; breastfeeding is safe; explain choose to disclose status if desired | 3.50 | 4.00 | 4.00 | 4.00 | 2 |

| Condition | Opening statement | Recommended treatment and counseling | Overall | Diagnosis | Social | Ease | *n* |
|---|---|---|---|---|---|---|---|
| HBV Not Eligible for Treatment | Doctor, I have had chronic hepatitis B for many years and I recently moved to this area, so I am going to be seeing you from now on. It has been about 6 months since my last exam. | Not eligible for treatment; no fibrosis or significant viral activity; no additional tests of liver function are necessary; mention follow up every 6-12 months for clinical exam, LFTs, fibrosis, HBV DNA, and HBeAg; explain disease is stable and no treatment needed; explain importance of regular monitoring; recommend screening family; recommend avoiding alcohol and live a healthy lifestyle | 3.00 | 3.00 | 3.00 | 3.00 | 1 |
| Hepatitis C | Doctor, I recently have felt very tired. My knees hurt, and I have had nausea. I went to a small clinic before coming here where they did some tests but my problems have not been resolved. | Outpatient treatment; eligible for DAA therapy: SOF/VEL or SOF+DAC for 12 weeks; after 1 month: LFTs, CBC, SVR12: HCV RNA, fibrosis, AFP, ultrasound, LFT, CBC; screen for HCC every 6 months after SVR 12 evaluation (as he has F3 fibrosis) | 4.00 | 4.00 | 4.00 | 4.00 | 2 |
| Hepatitis C with ARV | Doctor, I feel tired all the time, and my eyes have been a bit yellow since this happened. I am on antiretroviral therapy, is this related? | Continue ART (switch from TDF/3TC/EFV to TDF/3TC/DTG (TLD) regimen); begin SOF + DAC x 24 weeks; 4-week follow-up: LFTs, CBC, creatinine; SVR12 testing after 12 weeks post-treatment; continue HCC screening every 6 months; explain disease status and treatment benefits; explain importance of adherence to ARV and DAA; recommend monitoring of interactions and side effects; encourage partner and family testing; emphasize a healthy lifestyle and prevention of reinfection | 4.00 | 4.00 | 3.50 | 3.50 | 2 |

**Table S2. Summary of provider performance by case.** *N* is the number of coded and rated interactions for that case and Items is the number of checklist items in that case; in the overall row, Items is instead the rubric-item observations pooled across cases. Checklist completion is the pooled item-level proportion. Correct diagnosis is the proportion of interactions in which the provider named the condition, which scores all ten cases the same way. Qualifier identified is the proportion in which the provider also gave the stage or qualification that case's rule specifies (reproduced in Table S3); only the five hepatitis rules specify one, so the five general cases report no value. Duration is the time from the start of the conversation to the treatment decision, reported as median (IQR) because sessions were self-administered in a browser and could be left open. Intervals are provider-cluster bootstrap percentiles for checklist completion and for the pooled diagnosis and qualifier proportions in the overall row, and Clopper-Pearson exact for the per-case diagnosis and qualifier proportions, all at the 95% level.

| Condition | *N* | Items | Checklist (%) | Correct diagnosis (%) | Qualifier identified (%) | Duration (min) |
|---|---|---|---|---|---|---|
| Asthma | 14 | 62 | 12.1 ( 8.6, 15.8) | 64.3 (35.1, 87.2) | – | 8.4 ( 5.5, 9.1) |
| Pneumonia | 14 | 48 | 23.6 (20.1, 27.6) | 92.9 (66.1, 99.8) | – | 9.5 ( 5.0, 12.6) |
| Type II Diabetes Mellitus | 13 | 58 | 15.4 (10.7, 20.4) | 84.6 (54.6, 98.1) | – | 6.0 ( 4.4, 10.9) |
| Pulmonary Tuberculosis | 13 | 58 | 21.5 (17.3, 25.7) | 92.3 (64.0, 99.8) | – | 8.4 ( 5.5, 18.3) |
| Hypertension | 11 | 56 | 16.7 (13.1, 20.8) | 81.8 (48.2, 97.7) | – | 9.9 ( 6.3, 14.4) |
| Hepatitis B with Cirrhosis | 14 | 74 | 11.4 ( 7.1, 16.1) | 92.9 (66.1, 99.8) | 42.9 (17.7, 71.1) | 13.0 ( 7.7, 23.3) |
| Hepatitis B in Pregnancy | 12 | 61 | 13.3 ( 8.6, 18.2) | 91.7 (61.5, 99.8) | 16.7 ( 2.1, 48.4) | 9.6 ( 6.0, 16.8) |
| HBV Not Eligible for Treatment | 13 | 58 | 15.5 (12.1, 19.4) | 100.0 (75.3, 100.0) | 0.0 ( 0.0, 24.7) | 7.6 ( 4.3, 12.6) |
| Hepatitis C | 14 | 62 | 17.5 (12.6, 22.5) | 92.9 (66.1, 99.8) | 14.3 ( 1.8, 42.8) | 12.3 ( 5.9, 26.1) |
| Hepatitis C with ARV | 12 | 63 | 16.6 (11.5, 22.0) | 83.3 (51.6, 97.9) | 33.3 ( 9.9, 65.1) | 8.4 ( 4.7, 14.7) |
| *Overall* | 130 | 7814 | 16.1 (13.3, 18.8) | 87.7 (79.7, 93.9) | 21.5 (12.1, 30.8) | 9.0 ( 5.4, 14.7) |

**Table S3. Case-specific diagnosis scoring rules.** Each case contains exactly one diagnosis item. The second column reproduces that item's text verbatim from the scoring rubric, including the short item code the rubric uses internally. Each of the five hepatitis cases asks in addition for a stage or a qualification, and the Hepatitis B in Pregnancy and HBV Not Eligible for Treatment cases share the same rule text.

| Condition | Diagnosis rule as written in the rubric |
|---|---|
| Asthma | AD: correctly diagnose asthma? |
| Pneumonia | PD: correctly diagnose pneumonia? |
| Type II Diabetes Mellitus | DD: correctly diagnose diabetes? |
| Pulmonary Tuberculosis | TD:correctly diagnose TB? |
| Hypertension | HD: correctly diagnose hypertension? |
| Hepatitis B with Cirrhosis | HBCD: correctly diagnose hepatitis B with decompensated cirrhosis (Child-Pugh Class C)? |
| Hepatitis B in Pregnancy | HBPD: correctly diagnose chronic hepatitis B without fibrosis? |
| HBV Not Eligible for Treatment | HBND: correctly diagnose chronic hepatitis B without fibrosis? |
| Hepatitis C | HCD: correctly diagnose Chronic HCV with advanced fibrosis (F3), without cirrhosis and without co-infections? |
| Hepatitis C with ARV | HCAD: correctly diagnose Chronic viremic HCV infection, co-infected with HBV and HIV, compensated liver cirrhosis? |

### 3 Focus group supplementary results

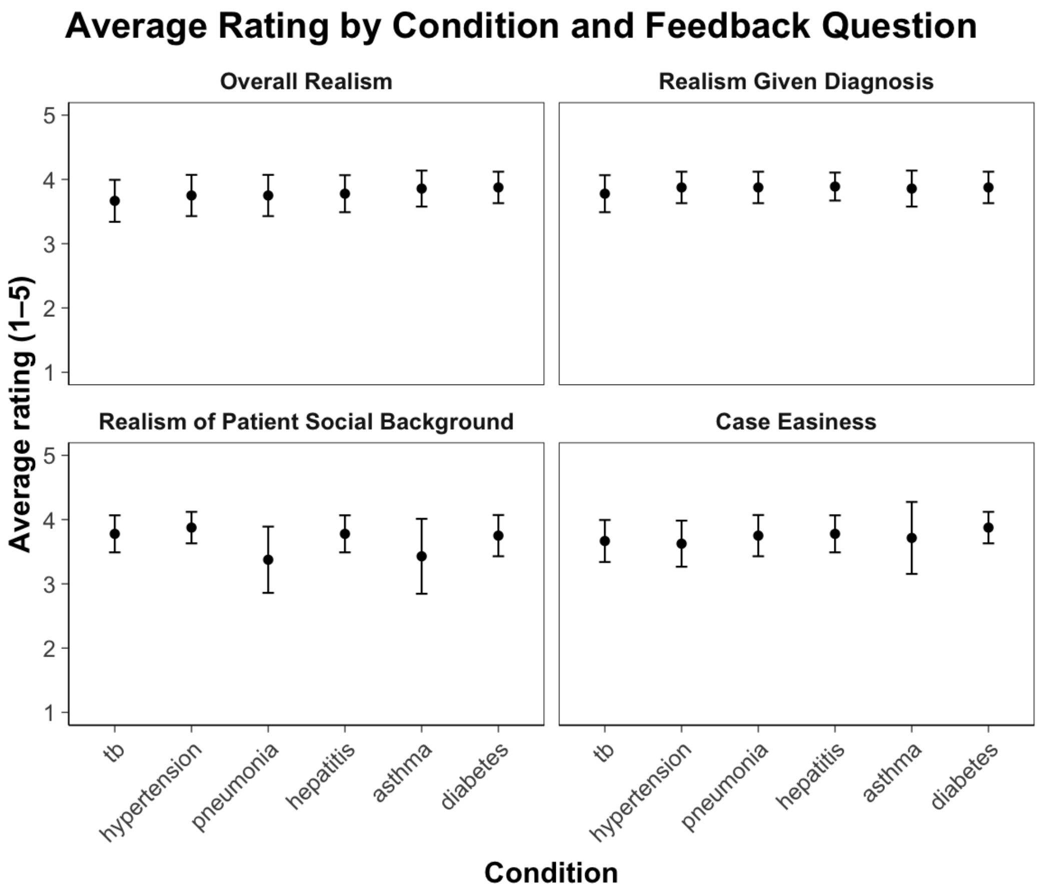


**Fig S1. Average realism rating by condition and feedback question.** Mean ratings (1–5 scale) with 95% confidence intervals are shown for each of six conditions completed in the focus group, across four feedback dimensions: overall realism, realism given diagnosis, realism of patient social background, and case easiness.

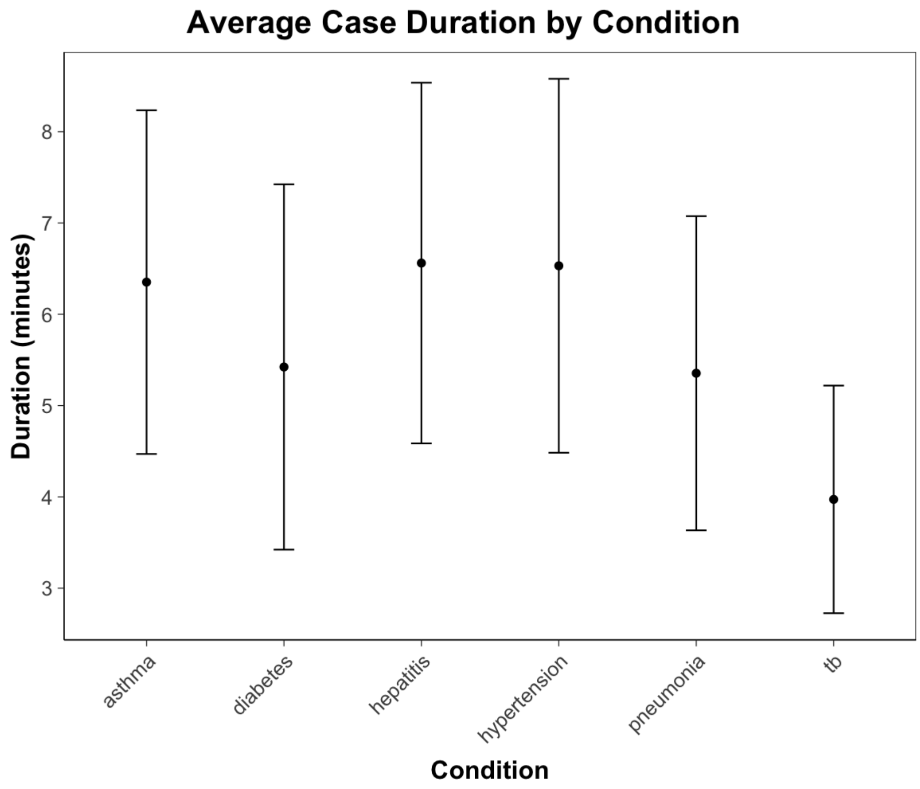


**Fig S2. Average case duration by condition in the focus group.** Mean duration in minutes with 95% confidence intervals for each condition.

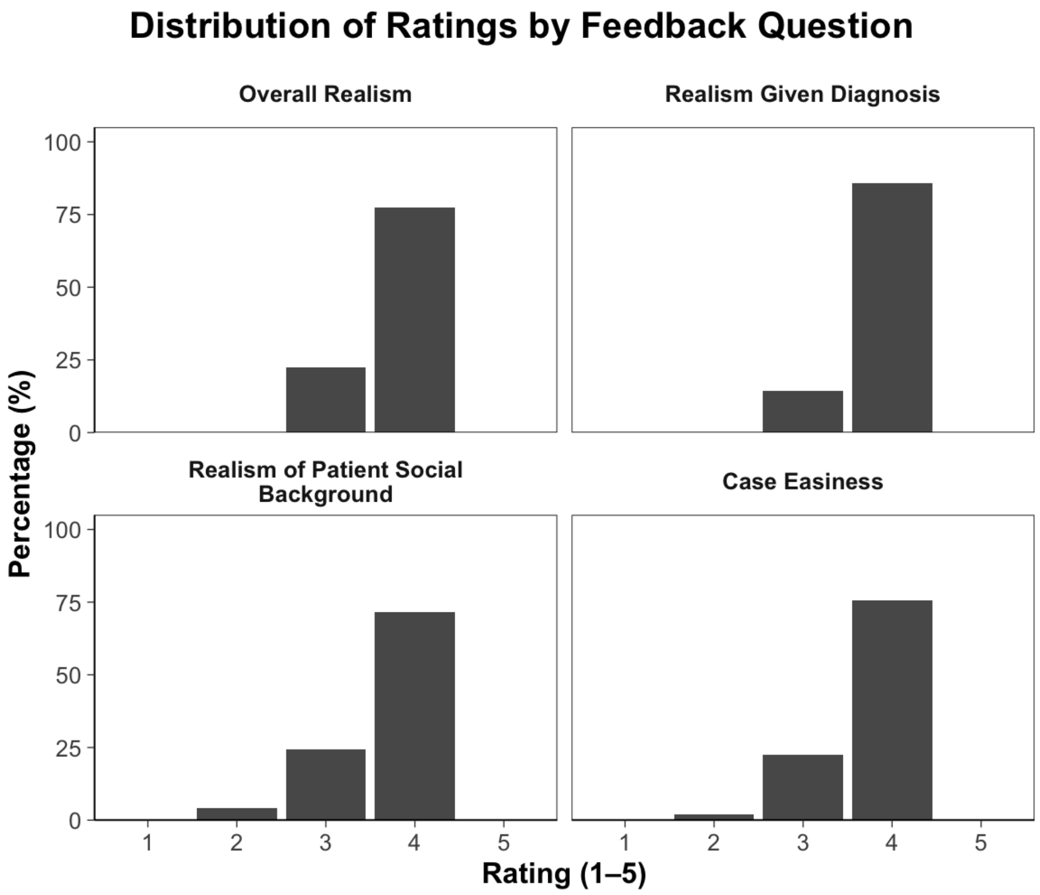


**Fig S3. Distribution of focus group realism ratings by feedback question.** Percentage of all ratings at each level (1–5) across four dimensions.

### 4 Automated scoring prompt

The classification script sent one prompt per interaction, reproduced below exactly as the script builds it. Braces mark the values substituted at run time: the transcript, the provider's stated diagnosis and treatment plan, the rubric items for that case serialized as JSON, the language of the transcript, and the number of rubric items the response must contain. Doubled braces are literal braces in the JSON response template.

```
You are evaluating a healthcare provider's clinical consultation transcript.
Determine whether the PROVIDER addressed each rubric item.

The transcript is in {lang_label}. Turns are labelled: `User:` is the PROVIDER
being assessed. `LLM:` is the simulated patient, and any nurse or laboratory
result reported under `LLM:` is also NOT the provider.

## CONSULTATION CONTEXT

**Transcript:**
{transcript}

**Provider's Diagnosis:** {diagnosis}

**Provider's Treatment Plan:** {treatment}

## RUBRIC ITEMS

{items_json}

Each item has an "action" ("ask about", "examine", "prescribe", ...). The
action describes something the PROVIDER must do.

## SCORING RULES

1. Score 1 only if the PROVIDER performed the action, evidenced by the
   provider's own words in a `User:` turn.
2. A patient volunteering information does NOT complete an "ask about" item.
   The provider must have asked. If the only support is a patient or nurse
   turn, score 0.
3. Laboratory or imaging results appearing in an `LLM:` turn do not by
   themselves complete an "order tests of" item. The provider must have
   ordered the test.
4. Quote verbatim. Do not paraphrase, and do not quote the rubric back.
5. If you score 1, set "speaker" to who said the quoted words: "provider" for
   a `User:` turn, "patient" for an `LLM:` turn. If the quote is not from the
   provider, reconsider the score under rules 2 and 3.
6. "confidence" is your probability from 0.0 to 1.0 that the score is correct.
7. If you score 0, set "evidence" and "speaker" to null and still give a
   confidence.

## RESPONSE FORMAT

Return a JSON object, no other text:

{{"results": [
  {{"id": "item_id", "score": 0, "confidence": 0.9, "evidence": null, "speaker": null}},
  {{"id": "item_id", "score": 1, "confidence": 0.8, "evidence": "verbatim quote", "speaker": "provider"}}
]}}

"results" must contain exactly {len(items)} entries, one per rubric item, in
the order given above. Never omit an item.
```

### 5 Chatbot fidelity audit prompt

The first block is the system message and the second is the user message. Braces mark the values substituted at run time: the case label, the scripted opening statement, the tests the case script expects, the correct management, and the transcript with its turns numbered and labelled. Doubled braces are literal braces in the JSON response template.

```
You audit whether a simulated-patient chatbot followed its clinical case
script. You never assert anything you cannot quote verbatim from the
transcript. You return only valid JSON.
```

```
Audit one consultation transcript for SIMULATED-PATIENT FIDELITY. You are not
scoring the provider. You are checking whether the chatbot playing the patient,
the nurse and the laboratory behaved as its case script requires.

## CASE: {case_label}

Scripted opening statement: {opening}

Tests the script expects a competent provider to order (for reference only --
the provider is free to order others, and failing to order one of these is the
PROVIDER's omission, not the chatbot's):
{items}

Correct management per the script: {treatment}

## TRANSCRIPT

Turns are numbered [T1], [T2], ... `User` is the PROVIDER. `LLM` is the
chatbot, speaking as the patient, as a nurse, or as the laboratory.

{turns}

## WHAT TO EXTRACT

### 1. orders
Every laboratory test, imaging study or bedside measurement the PROVIDER
requested in a `User` turn.

  - One entry per NAMED test or NAMED panel. "blood count, creatinine and
    chest X-ray" is three entries. "hepatitis tests" is one entry, as is
    "liver function tests" -- a named panel is one order, not one per
    constituent.
  - A vague request naming nothing ("I will run some tests", "I will give you
    the necessary tests") is ONE entry with test "unspecified tests".
  - Only the provider can order. A nurse offering a test is not an order.
  - If the provider orders the same test twice, make one entry, order_turn =
    the first request, and set reordered true.

For each order set "outcome" to exactly one of:
  value_returned          a later LLM turn gives the actual value or finding
  explicitly_unavailable  a later LLM turn says the result is not available,
                          not done, or pending
  promised_never_delivered  an LLM turn promises the result ("I will report it
                          as soon as it is ready") and no value ever follows
  acknowledged_only       an LLM turn acknowledges the order and nothing
                          further is ever said about it
  no_response             no LLM turn ever mentions this test again

A promise, an acknowledgement, or a restatement of what was ordered is NOT a
result. Only an actual value, measurement or descriptive finding counts.

Be generous about wording and be strict only about substance. A test counts as
returned when the value is there under any name: HGB is haemoglobin, GOT is
AST, GPT is ALT, ARN is RNA, abdominal ultrasound covers liver ultrasound,
```

"blood count" is answered by HGB and platelets, "hepatitis screening" is answered by HBsAg / HBV DNA / anti-HCV / HCV RNA, "liver function" is answered by AST, ALT, bilirubin and albumin, and "blood biochemistry: creatinine 80" returns creatinine. A test ordered twice and returned once is returned. Mark an order unreturned only when nothing in any later LLM turn gives its value.

##### 2. unrequested_results
Results the chatbot returned for a specific test the provider never named individually. Quote the result line. Set "covered_by_broader_order" true when the test still falls inside something the provider did ask for -- a vague "some tests", or a panel such as "blood count" or "hepatitis tests" -- and false when the provider asked for nothing that covers it at all. Both are the chatbot choosing what laboratory work to report; the flag separates the chatbot expanding a request from the chatbot inventing one.

##### 3. volunteered_history
LLM turns where the patient discloses clinically relevant information the provider did not ask for in the immediately preceding provider turn. The scripted opening statement itself is NOT volunteered information. Answering the question asked is not volunteering. Adding a symptom, exposure, history or family history nobody asked about is.

##### 4. contradictions
Two LLM turns that report incompatible facts about the same patient (a value that changes, a symptom denied then affirmed, an arithmetic result that does not follow from the values given). Quote both.

##### 5. character_breaks
The chatbot stepping outside the simulation: speaking as the doctor, giving the diagnosis or management plan the provider was supposed to give, referring to itself as a model or assistant, or discussing the exercise as an exercise.

#### RULES

Every "quote" field must be copied verbatim from the transcript, long enough to locate (at least 8 words where the source allows). Never paraphrase. Never quote this instruction back. If a list is empty, return [].

#### RESPONSE FORMAT

Return this JSON object and nothing else:

```
{{"orders": [{{"order_turn": 4, "test": "chest X-ray", "category": "imaging",
   "order_quote": "verbatim from the User turn", "reordered": false,
   "outcome": "value_returned", "result_turn": 5,
   "result_quote": "verbatim from the LLM turn"}}],
 "unrequested_results": [{{"turn": 5, "test": "urinalysis", "quote": "verbatim",
   "covered_by_broader_order": false}}],
 "volunteered_history": [{{"turn": 3, "quote": "verbatim", "what": "short phrase"}}],
 "contradictions": [{{"turns": [5, 9], "quote_a": "verbatim", "quote_b": "verbatim",
   "what": "short phrase"}}],
 "character_breaks": [{{"turn": 7, "quote": "verbatim", "what": "short phrase"}}]}}
```

"category" is one of "laboratory", "imaging", "bedside", "other".
Set "result_turn" and "result_quote" to null when outcome is not "value_returned".

### 6 STROBE checklist for cross-sectional studies

**Study Design:** Cross-sectional validation study
**Study Registration:** Not registered; study is a methods paper validating an assessment tool

This checklist reproduces the 22 items of the STROBE Statement checklist for cross-sectional studies and names the manuscript section reporting each one. Item wording follows the published form, including its British spellings. Sections are named rather than paged because the pagination of a typeset article changes at every stage of review.

| Item | STROBE Checklist Item | Reported in |
|---|---|---|
| *Title and abstract* | | |
| 1 | (a) Indicate the study's design with a commonly used term in the title or the abstract<br>(b) Provide in the abstract an informative and balanced summary of what was done and what was found | Title; Abstract<br>Abstract |
| *Introduction* | | |
| 2 | **Background/rationale:** Explain the scientific background and rationale for the investigation being reported | Introduction, first and second paragraphs |
| 3 | **Objectives:** State specific objectives, including any prespecified hypotheses | Introduction, third paragraph; Materials and methods, "Outcomes". No hypothesis was prespecified and the study is not registered |
| *Methods* | | |
| 4 | **Study design:** Present key elements of study design early in the paper | Abstract; Introduction, third paragraph; Materials and methods, "Study design and participants" |
| 5 | **Setting:** Describe the setting, locations, and relevant dates, including periods of recruitment, exposure, follow-up, and data collection | Materials and methods, "Study design and participants" |
| 6 | **Participants:** Give the eligibility criteria, and the sources and methods of selection of participants | Materials and methods, "Study design and participants" |
| 7 | **Variables:** Clearly define all outcomes, exposures, predictors, potential confounders, and effect modifiers. Give diagnostic criteria, if applicable | Materials and methods, "Data extraction and coding" and "Outcomes" |
| 8 | **Data sources/measurement:** For each variable of interest, give sources of data and details of methods of assessment (measurement). Describe comparability of assessment methods if there is more than one group | Materials and methods, "Chatbot implementation", "Data extraction and coding" and "Chatbot fidelity audit" |
| 9 | **Bias:** Describe any efforts to address potential sources of bias | Materials and methods, "Data extraction and coding" and "Statistical analysis", which report the single-coder reference standard, the independent and mutually blinded expert raters, and the confounding of rater with language; Discussion, "Limitations" |

*Continued on next page*

| Item | STROBE Checklist Item | Reported in |
|---|---|---|
| 10 | **Study size:** Explain how the study size was arrived at | Materials and methods, "Study design and participants". No target size was set for this pilot; the resulting imprecision is stated in Discussion, "Limitations" |
| 11 | **Quantitative variables:** Explain how quantitative variables were handled in the analyses. If applicable, describe which groupings were chosen and why | Materials and methods, "Data extraction and coding" and "Statistical analysis" |
| 12 | (a) Describe all statistical methods, including those used to control for confounding | Materials and methods, "Statistical analysis": Pearson correlations and OLS regressions estimated on the pooled sample with standard errors clustered at the provider level, ROC and AUROC analysis, Clopper-Pearson exact intervals, and a provider-cluster bootstrap |
| | (b) Describe any methods used to examine subgroups and interactions | Materials and methods, "Statistical analysis". Session-level estimates are pooled across scenarios rather than computed within them; agreement is reported separately by rubric domain and by coding language in Results, "Validation sample" |
| | (c) Explain how missing data were addressed | Materials and methods, "Study design and participants". Interactions that were neither coded nor expert-rated are excluded from the session-level and item-level analyses rather than imputed |
| | (d) If applicable, describe analytical methods taking account of sampling strategy | Materials and methods, "Study design and participants" and "Statistical analysis". Each provider was assigned six of the ten cases at random, and standard errors are clustered on the provider |
| | (e) Describe any sensitivity analyses | Results, "Validation sample". The reverse-direction regression is reported as a check on scale compression |
| *Results* | | |
| 13 | (a) **Participants:** Report numbers of individuals at each stage of study: numbers potentially eligible, examined for eligibility, confirmed eligible, included in the study, completing follow-up, and analysed | Materials and methods, "Study design and participants"; Results, "Focus group" and "Validation sample" |
| | (b) Give reasons for non-participation at each stage | Results, "Focus group" and "Validation sample". Reasons for non-completion were not recorded |
| | (c) Consider use of a flow diagram | Not applicable; a single consented cohort with no screening stages |

*Continued on next page*

| Item | STROBE Checklist Item | Reported in |
|---|---|---|
| 14 | (a) **Descriptive data:** Give characteristics of study participants (eg demographic, clinical, social) and information on exposures and potential confounders | Materials and methods, "Study design and participants". No individual provider characteristics were collected beyond the recruiting site and training program |
| | (b) Indicate number of participants with missing data for each variable of interest | Materials and methods, "Study design and participants": 2 of the 132 fielded interactions were neither coded nor expert-rated |
| 15 | **Outcome data:** Report numbers of outcome events or summary measures | Results, "Validation sample" and "Chatbot fidelity" |
| 16 | (a) **Main results:** Give unadjusted estimates and, if applicable, confounder-adjusted estimates and their precision (eg, 95% confidence interval). Make clear which confounders were adjusted for and why they were included | Results, "Validation sample": Pearson correlations, OLS slopes and $R^2$, and AUROCs, each with a 95% confidence interval. No confounder adjustment is applied |
| | (b) Report category boundaries when continuous variables were categorized | Materials and methods, "Data extraction and coding", where checklist items are binary, and "Statistical analysis", where expert ratings run on a 1–5 scale averaged within rater |
| | (c) If relevant, consider translating estimates of relative risk into absolute risk for a meaningful time period | Not applicable; no relative risk is estimated |
| 17 | **Other analyses:** Report other analyses done: analyses of subgroups and interactions, and sensitivity analyses | Results, "Validation sample", which reports agreement by rubric domain and the English against Vietnamese comparison, and "Chatbot fidelity" |
| *Discussion* | | |
| 18 | **Key results:** Summarise key results with reference to study objectives | Discussion, first paragraph |
| 19 | **Limitations:** Discuss limitations of the study, taking into account sources of potential bias or imprecision. Discuss both direction and magnitude of any potential bias | Discussion, "Limitations" |
| 20 | **Interpretation:** Give a cautious overall interpretation of results considering objectives, limitations, multiplicity of analyses, results from similar studies, and other relevant evidence | Discussion, first to third paragraphs |
| 21 | **Generalisability:** Discuss the generalisability (external validity) of the study results | Discussion, second and third paragraphs; "Limitations" and "Conclusion" |
| *Other information* | | |
| 22 | **Funding:** Give the source of funding and the role of the funders for the present study and, if applicable, for the original study on which the present article is based | Financial disclosure |
